# Persistence of SARS-CoV-2 immunity, Omicron’s footprints, and projections of epidemic resurgences in South African population cohorts

**DOI:** 10.1101/2022.02.11.22270854

**Authors:** Kaiyuan Sun, Stefano Tempia, Jackie Kleynhans, Anne von Gottberg, Meredith L McMorrow, Nicole Wolter, Jinal N. Bhiman, Jocelyn Moyes, Mignon du Plessis, Maimuna Carrim, Amelia Buys, Neil A Martinson, Kathleen Kahn, Stephen Tollman, Limakatso Lebina, Floidy Wafawanaka, Jacques D. du Toit, Francesc Xavier Gómez-Olivé, Thulisa Mkhencele, Cécile Viboud, Cheryl Cohen, the PHIRST group

**Affiliations:** Division of International Epidemiology and Population Studies, Fogarty International Center, National Institutes of Health, Bethesda, Maryland, United States of America; Centre for Respiratory Diseases and Meningitis, National Institute for Communicable Diseases of the National Health Laboratory Service, Johannesburg, South Africa; School of Public Health, Faculty of Health Sciences, University of the Witwatersrand, Johannesburg, South Africa; Influenza Division, Centers for Disease Control and Prevention, Atlanta, Georgia, United States of America; School of Pathology, Faculty of Health Sciences, University of the Witwatersrand, Johannesburg, South Africa; Perinatal HIV Research Unit, University of the Witwatersrand, South Africa; Johns Hopkins University Center for TB Research, Baltimore, Maryland, United States of America; MRC/Wits Rural Public Health and Health Transitions Research Unit (Agincourt), School of Public Health, Faculty of Health Sciences, University of the Witwatersrand, Johannesburg, South Africa

## Abstract

Understanding the build-up of immunity with successive SARS-CoV-2 variants and the epidemiological conditions that favor rapidly expanding epidemics will facilitate future pandemic control. High-resolution infection and serology data from longitudinal household cohorts in South Africa reveal high cumulative infection rates and durable cross-protective immunity conferred by prior infection in the pre-Omicron era. Building on the cohort’s history of past exposures to different SARS-CoV-2 variants and vaccination, we use mathematical models to explore the fitness advantage of the Omicron variant and its epidemic trajectory. Modelling suggests the Omicron wave infected a large fraction of the population, leaving a complex landscape of population immunity primed and boosted with antigenically distinct variants. Future SARS-CoV-2 resurgences are likely under a range of scenarios of viral characteristics, population contacts, and residual cross-protection.

**One Sentence Summary:** Closely monitored population in South Africa reveal high cumulative infection rates and durable protection by prior infection against pre-Omicron variants. Modelling indicates that a large fraction of the population has been infected with Omicron; yet epidemic resurgences are plausible under a wide range of epidemiologic scenarios.

## Main Text

A key questions for the long-term control of SARS-CoV-2 is how sequential exposures to different variants of the virus shape population immunity and thereby modulate subsequent epidemic cycles and disease burden. Few studies have characterized the protection conferred by infection over long time periods, particularly in low and middle-income settings where vaccine access is limited and high SARS-CoV-2 infection rates have been reported *(1–4)*. Here we used unique data from two prospectively followed cohorts in South Africa to estimate the strength of cross-protective immunity conferred by infection with successive SARS-CoV-2 variants. We relied on the cohort data to reconstruct the landscape of population immunity prior to the emergence of the Omicron variant, and modeled the trajectory, scale and long-term consequences of the Omicron epidemic in this population.

South Africa experienced three distinct SARS-CoV-2 epidemic waves prior to the emergence of the Omicron variant, with the first wave (June 2020 – December 2020) dominated by the ancestral SARS-CoV-2 strain carrying the D614G mutation (refer to as D614G hereafter) *(5)*, the second wave (December 2020 – May 2021) dominated by the Beta (B.1.351) variant *(6)*, and the third wave (May 2021 – October 2021) dominated by the Delta (B.1.617.2) variant *(7)*. In South Africa, the Omicron variant was first identified in the province of Gauteng Province in November 2021, and swiftly spread nationally and globally, causing rapid growth in case counts relative to prior waves *(7, 8)*. Similar patterns of rapid growth despite high levels of pre-existing immunity from infection and vaccination have also been reported in numerous countries across the world, with Omicron replacing Delta in multiple global locations, even when prevalence of Delta was high *(9, 10)*.

The apparent fitness advantage of the Omicron variant over Delta could be driven by immune evasion, increased intrinsic transmissibility, or a combination of both. The immune evasion hypothesis is supported by an increased reinfection risk coinciding with the rise of the Omicron variant *(7, 8, 11, 12)*. Further support for this hypothesis comes from *in vitro* analyses of sera from convalescent patients (infected with pre-Omicron variants) and vaccinated individuals, which show reduced neutralization titers against Omicron compared to earlier variants *(13–16)*. Similarly, data from multiple settings have shown decreased vaccine effectiveness against Omicron *(17, 18)*. Separately, epidemiological and experimental data point to a reduced clinical severity of Omicron *(17, 19)*, possibly due to increased tropism for the upper respiratory tract rather than the lung, which could also promote higher transmission relative to pre-Omicron variants. As the Omicron wave subsides, the relative contribution of these factors to Omicron’s spread remains elusive, in part due to uncertainty in the level of population immunity before the rise of Omicron. Further, Omicron’s rapid spread poses immense pressure on SARS-CoV-2 testing capabilities, and its relatively benign course in most people make it difficult to assess the full scope of the epidemic.

To understand the long-term dynamics of SARS-CoV-2, we leveraged data from two longitudinal household cohorts followed over a 13-month period, from July 2020 to August 2021 in rural and urban areas of South Africa (**P**rospective **H**ousehold study of SARS-CoV-2, **I**nfluenza and **R**espiratory **S**yncytial virus community burden, **T**ransmission dynamics and viral interaction in South Africa, PHIRST-C, where “**C**” stands for COVID-19, previously described in *(4)*). We relied on the results of densely sampled respiratory and serologic specimens testing from 222 households to model the kinetics of viral shedding, transmission dynamics among household members, and cross-protection between successive variants circulating prior to the emergence of Omicron. We used population-level models calibrated against data from these prospective cohorts and surveillance efforts to clarify long-term patterns of immunity acquisition, the impact of immune evasion, and future epidemic trajectories for SARS-CoV-2 in the aftermath of the Omicron wave.

### Overview of SARS-CoV-2 epidemiology in study sites

The PHIRST-C cohort captured the dynamics of three waves of SARS-CoV-2 infections in rural and urban site located in two provinces of South Africa (see methods for details). In total, 1,200 individuals living in 222 randomly selected households were enrolled and followed up twice a week for SARS-CoV-2 real-time reverse transcription polymerase chain reaction (rRT-PCR) testing and symptom monitoring, while blood draws were obtained every 2 months for SARS-CoV-2 serologic tests. Throughout the study, we used a broad measurement of prior and ongoing infections including both serologic and virologic evidence, irrespective of symptoms. In Figure 1A-B, we show the weekly SARS-CoV-2 epidemic curves in the district where each study site was located *(20)*. Population vaccination started in June 2021 in South Africa and the fraction of the cohort population fully vaccinated remained below 10% at the conclusion of the study in September 2021 (Figure 1A-B). Both Pfizer/BioNTech’s BNT162b2 and J&J/Janssen’s Ad26.COV2.S vaccines are used in South Africa (Figure S1). We did not evaluate vaccine effectiveness in this study and focused on protection conferred by prior SARS-CoV-2 infection. However, we consider the impact of vaccination in projections of the Omicron wave and post-Omicron future.

**Fig. 1.**
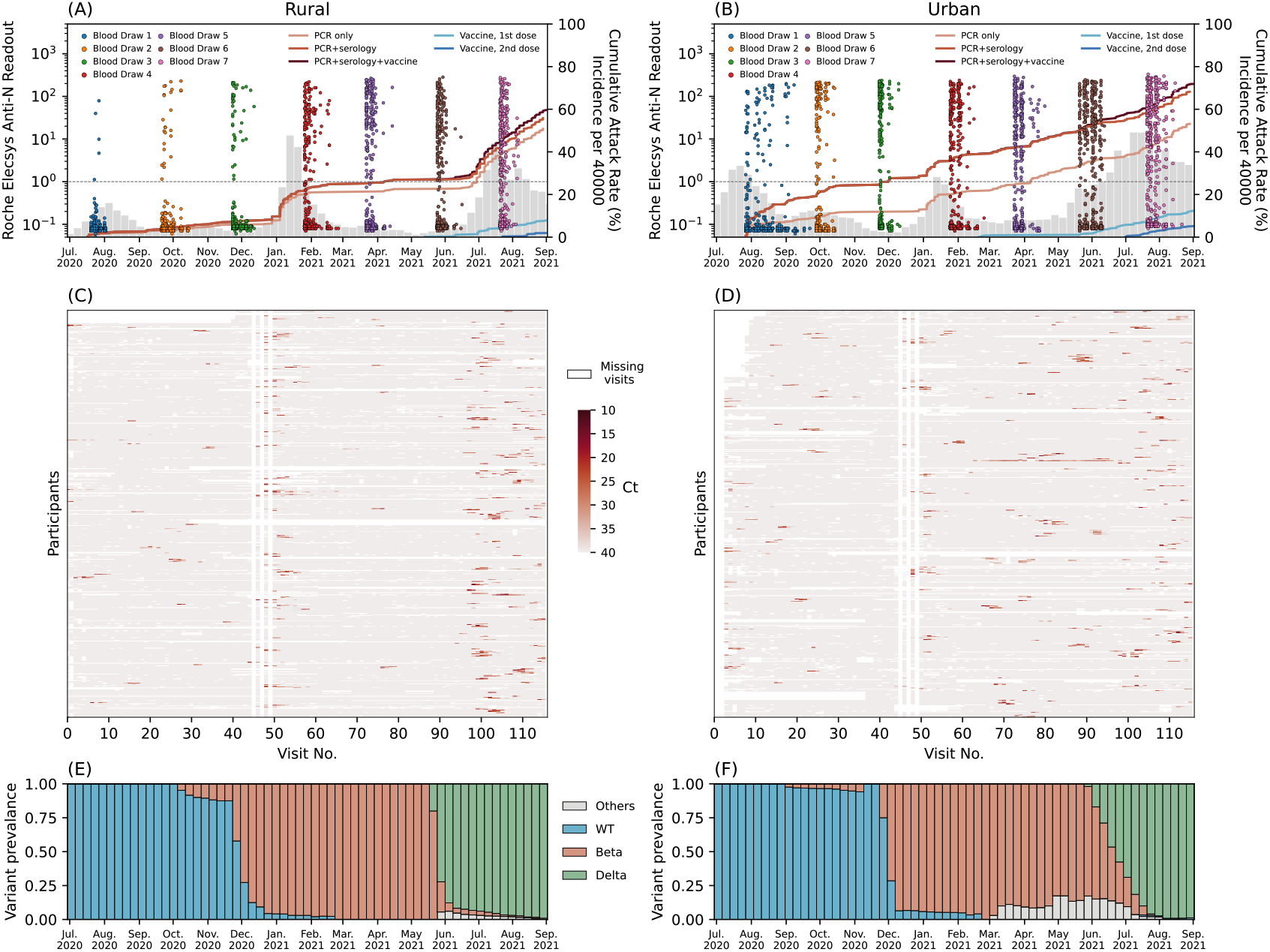
PHIRST-C study June 2020 – September 2021, description of the epidemiology of SARS-CoV-2 in the two study sites, along with serology and rRT-PCR data. (A) Dots in different colors represent the timing and the readouts (axis on the left) of Roche Elecsys Anti-SARS-CoV-2 assay of serum specimens collected from 4 different blood draws of the rural cohort. The dash line is the positive cutoff of the Roche Elecsys Anti-SARS-CoV-2 assay, above which a specimen is considered sero-positive. The red lines (from light to dark) are the cumulative SARS-CoV-2 variant exposures (axis on the right) over time, captured by positive rRT-PCR of mid-turbinate nasal swab samples only; by either positive serum antibody or positive mid-turbinate nasal swabs by rRT-PCR, and by either positive serum antibody or positive mid-turbinate nasal swabs by rRT-PCR or at least one dose of vaccine. The light and dark blue lines are the cumulative fraction of population receiving a 1^st^ and 2^nd^ dose of vaccine. The grey bars are the weekly SARS-CoV-2 incidence per 40,000 population (sharing the same axis on the right) captured by the surveillance system of Ehlanzeni District in Mpumalanga Province, where the rural site is located. (B) Same as (A) but for the urban site of Klerksdorp in the Dr Kenneth Kaunda District, North West Province. (C) rRT-PCR test results for all mid-turbinate nasal specimens collected from individuals in the rural cohort over 80 visits during the 13-month study period. Color white indicates missing specimens; color red indicates the Ct value of the rRT-PCR test, the darker the red color, the lower the Ct value. (D) Same as (C) but for the urban cohort. (E) The bi-monthly relative prevalence of D614G mutation, Beta, Delta and other variants over time at the rural site. (F) Same as (E) but for the urban site.

In the rural site, baseline enrollment visits started prior to the peak of the first epidemic wave. The seroprevalence of anti-SARS-CoV-2 nucleocapsid antibodies was 1.1% (5/445) at enrollment, increased to 7.3% (42/574) after the first wave (third blood draw), 25.4% (151/595) after the second wave (fifth blood draw), and reached 39.1% (227/581) around the peak of the third wave (seventh blood draw). The timing and individual results of the serological assay are visualized in Figure 1A. During the study period (July 2020 to August 2021), 50.9% (327/643) of individuals tested positive by rRT-PCR for at least one infection episode. The cumulative infection rate (confirmed by either a rRT-PCR or a serological test) was 59.7% (384/643) by the end of the study in the rural cohort. In contrast, in the urban site, enrollment started near the time of the peak of the first wave and SARS-CoV-2 seroprevalence was higher at enrollment (14.3%, 73/511), increased to 27.0% (143/530) after the first wave, 40.3% (207/514) after the second wave, and reached 55.7% (279/501) around the peak of the third wave (see Figure 1B for bi-monthly results). During the study period, 53.1% (296/557) of participants tested positive by rRT-PCR for at least one infection episode. The cumulative infection rate (confirmed by either a rRT-PCR or a serological test) was 69.4% (387/557) in the urban cohort by the end of the study.

In total across both sites, we observed 669 rRT-PCR-confirmed infection episodes, including 599/669 (89.5%) primary infections and 70/669 (9.5%) reinfections. The weekly incidence of SARS-CoV-2 infections within each cohort (Figure S2) matches the epidemic trajectory at the district level (Figure 1A-B), except for a less pronounced third wave in the urban cohort compared to that of the district. The Lineage-specific rRT-PCR and sequencing data revealed that 14.3% (96/669) of infections were D614G, 33.2% (222/669) were Beta, 44.1% (295/669) were Delta, 2.7% (18/669) were other lineages including Alpha and C.1.2 variants, and 5.7% (38/669) were inconclusive. Figure 1E-F shows the relative prevalence of different lineages over time for the rural and urban site, respectively (details in Materials and Methods Section 3.2).

### Kinetics of viral RNA shedding

To study the risk of infection and re-infection in the cohort, and better understand acquisition of immunity before the rise of Omicron, we first built a time-varying model that captured the dynamics of viral RNA shedding for each individual in the cohort, adjusted for host characteristics and variant types. Household exposure depends on the level of viral shedding among household members; to obtain a correlate of shedding intensity, we used the serial Ct values of nasal swab specimens collected twice-weekly and tested for SARS-CoV-2 using rRT-PCR. We considered Ct value of the rRT-PCR test of a specimen as a proxy for the RNA shedding intensity. We used the serial rRT-PCR test results to model the shedding kinetics of SARS-CoV-2 infection episodes, following prior work *(21, 22)*. To account for the potential role of adaptive immunity in limiting transmission in the later phases of infection, we allowed for different transmission risks during the viral RNA proliferation stage (before peak shedding) and the viral RNA clearance stage (after peak shedding), with shedding increases and decreases assumed to following linear curves on the scale of Ct values. Because the nasal swab sampling period ended on August 28, 2021, around the peak of the Delta wave in both sites, we limit our analysis to infection episodes with first positive PCR specimen 30 days prior to the end of sampling to avoid censoring bias.

Figure 2 A-C shows the RNA shedding kinetics of the D614G, Beta, and Delta variants respectively. All three variants had similar shedding kinetics characterized by a short proliferation stage (Figure 2E, median and interquartile range (IQR) for D614G: 3.2 (2.1 – 4.0) days, Beta: 3.3 (2.2 – 4.3) days, Delta: 3.1 (2.0 – 3.8) days) and a longer clearance stage (Figure 2F, median and IQR for D614G: 7.4 (4.3 – 10.2) days, Beta: 7.5 (5.0 – 9.3) days, Delta: 8.0 (5.7 – 9.5) days). The symptomatic rates among infection episodes were low across all variants, at 13% for D614G, 16% for Beta, and 18% for Delta (Figure 2A-C). The timing of symptom onset coincided with the timing of peak viral shedding (Figure 2A-C), suggesting significant shedding had already occurred prior to symptom presentation. After adjusting for age, sex, body mass index (BMI), and HIV infection status, symptomatic infections had significantly lower trough *Ct* value (peak shedding intensity) than asymptomatic infections (Figure 3A). Beta variant’s trough *Ct* value was lower than D614G, while Delta’s was the lowest among the three (Figure 3A). We also found that prior infection significantly reduced peak shedding by 4.0 Ct (95% CI 2.3 – 5.7) and shedding duration by 3.3 days (95% CI 1.9 – 4.7) upon reinfection (Figure 3A-B).

**Fig. 2.**
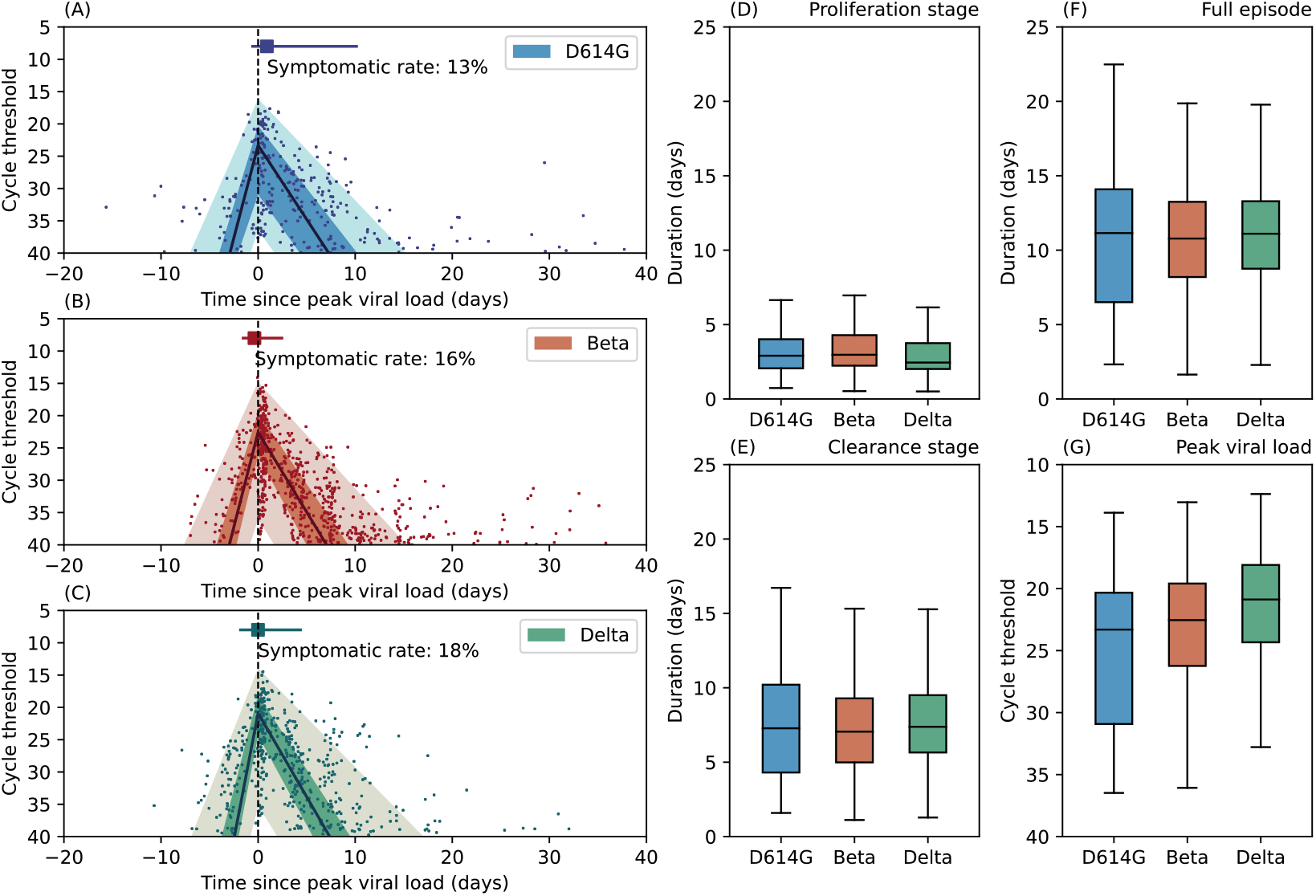
(A) Characterization of the RNA shedding kinetics for D614G infections. The solid dots are longitudinal Ct values observation for each infection episode, aligned based on the estimated timing of trough *Ct*. The solid line is the population median of all individual fits, the dark shade is the interquartile range, and the light shade is the 95% confidence interval. Dashed vertical line indicate the timing of peak viral load. The square marker and the horizontal line indicate the median time and interquartile range of symptom onset for symptomatic infections. We also reported the fraction of symptomatic infections among all infections (symptomatic rate) for D614G. (B) Same as (A) but for Beta variant. (C) Same as (A) but for Delta variant. Distribution of the estimated duration of the viral RNA proliferation stage. (D) Distribution of the estimated duration of the viral RNA proliferation stage for D614G, Beta, and Delta variants. Boxplots show median, interquartile range, minimum and maximum of the distribution (E) Same as (D) but for the distribution of the estimated duration of the viral RNA clearance stage. (F) Same as (D) but for the distribution of the estimated full duration (proliferation stage + clearance stage) of rRT-PCR positivity. (G) Same as (D) but for the distribution of the estimated peak *dCt*.

**Fig. 3.**
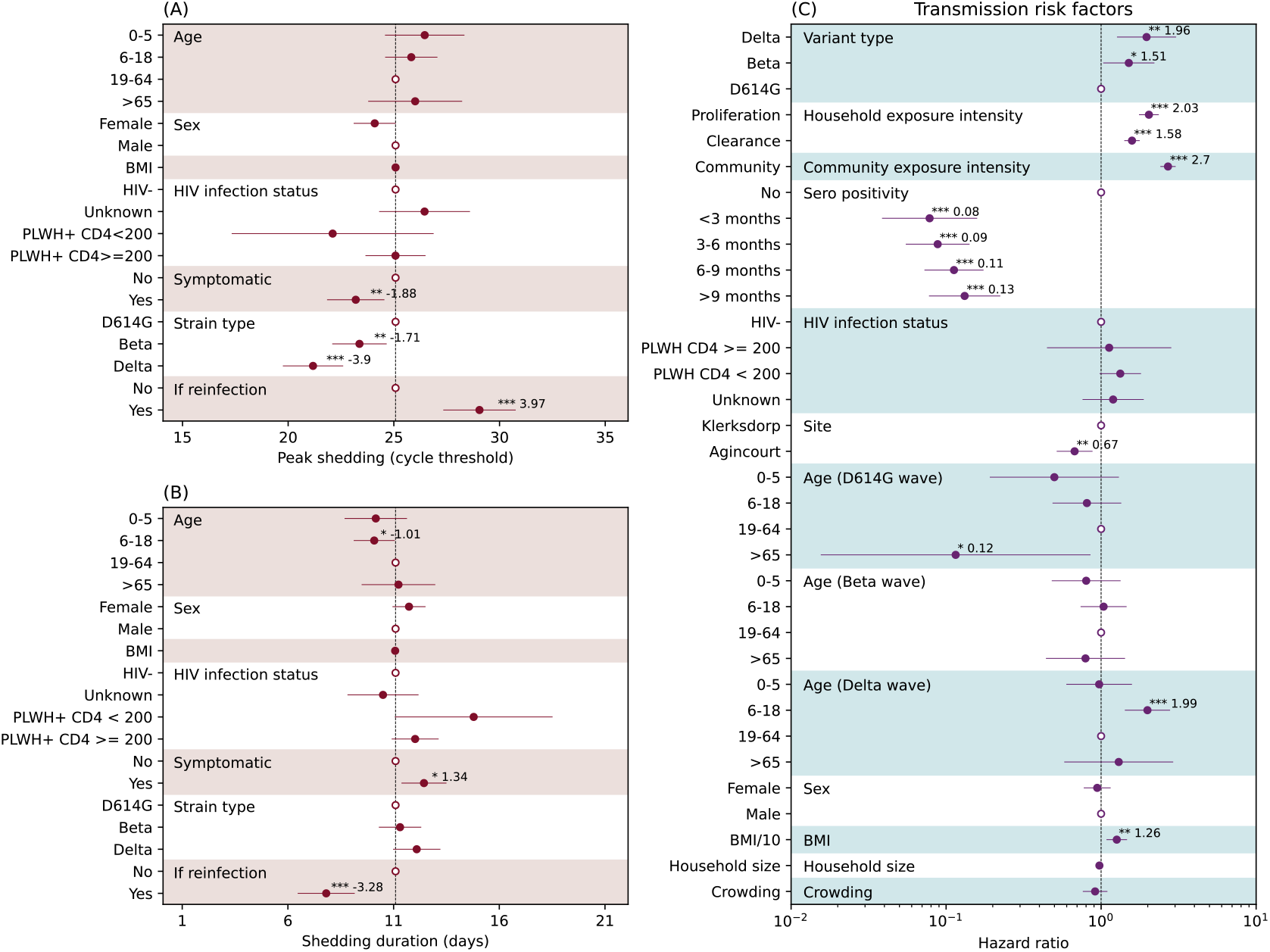
(A) The association between peak shedding (trough Ct) and age, sex BMI, HIV infection status, symptom presentation, variant type, and prior infection history, based on gaussian multiple regression. Regression coefficients along with 95%CIs are reported as solid dots and horizontal lines relative to the value of the regression intercept. The hollow dots are reference class for each of the categorical variable. (B) Same as (A) but for shedding duration. (C) Piecewise exponential hazard model on risk factors associated with infection acquisition. Hazard ratios (HR) along with 95%CIs are reported as solid dots and horizontal lines. The hollow dots are reference class for each of the categorical variable. Protection is measured as 1 − *HR*. *Indicates p<0.05; ** indicates p<0.01; *** indicates p < 0.001. Abbreviations: HIV- (HIV-uninfected individuals), PLWH+ CD4 <200 (Persons living with HIV, CD4+ T cell count under 200 cells/ml), PLWH+ CD4 >=200 (Persons living with HIV, CD4+ T cell count equal or above 200 cells/ml).

The population of the PHIRST-C cohort had a high prevalence of HIV, 13% in the rural site and 16% in the urban site, reflecting the burden of HIV infections in South Africa (Table 1). However, in this cohort, most (93.8%) persons living with HIV (PLWH) had CD4+ T cell counts ≥ 200 cells/ml (Table 1), and they did not differ from HIV-uninfected individuals in terms of SARS-CoV-2 shedding (Figure 3A-B). We found that individuals with low CD4+ T cell counts (< 200 cells/ml) tended to shed for longer and with lower trough *Ct* when compared to HIV-uninfected individuals but there was limited statistical power to detect these differences (Figure 3A-B).

**Table 1:** PHIRST-C study June 2020 – August 2021, characteristics of the population and SARS-CoV-2 infections at two study sites, South Africa. *PLWH: persons living with HIV

|  | **Rural** | **Urban** |
| --- | --- | --- |
| **Characteristics** | No. (%) | No. (%) |
| All | 643 (100) | 557 (100) |
| **Age group, in years** |  |  |
| 0-5 | 99 (15) | 55 (10) |
| 6-18 | 299 (47) | 211 (38) |
| 19-65 | 218 (34) | 260 (47) |
| >65 | 27 (4) | 31 (5) |
| **Sex** |  |  |
| Male | 234 (36) | 249 (45) |
| Female | 409 (64) | 308 (55) |
| **Household size** |  |  |
| <4 | 44 (7) | 62 (11) |
| 4-6 | 282 (44) | 296 (53) |
| 7-10 | 257 (40) | 141 (25) |
| >10 | 60 (9) | 58 (10) |
| **BMI** |  |  |
| Underweight | 259 (40) | 140 (25) |
| Healthy weight | 206 (32) | 192 (35) |
| Overweight | 93 (14) | 103 (18) |
| Obesity | 85 (13) | 120 (22) |
| Unknown | 0 (0) | 2 (0) |
| **HIV status** |  |  |
| Negative | 520 (81) | 451 (81) |
| PLWH*: CD4 < 200 | 5 (1) | 9 (2) |
| PLWH*: CD4 ≥ 200 | 75 (12) | 78 (14) |
| Unknown | 43 (7) | 19 (3) |
| **Vaccination status** |  |  |
| None | 593 (92) | 488 (88) |
| J&J | 12 (2) | 9 (2) |
| Pfizer first dose only | 27 (4) | 32 (6) |
| Pfizer first and second doses | 11 (2) | 28 (5) |

### Infection risk and protection against reinfection

Reconstruction of the variant-specific shedding kinetics of each infected individual allowed us not only to infer the timing of their infections, but also to evaluate the daily intensity of exposure to SARS-CoV-2 within their households. We used a piecewise exponential hazard model (detailed in Materials and Methods Section 3.5) to explore how variant type and prior infection history affect the risk of SARS-CoV-2 infection and reinfection, after adjustment for time-varying SARS-CoV-2 exposure and host factors. We regressed the daily infection risk of each individual on covariates including variant type, time since prior SARS-CoV-2 infection, age (allowing for variant-specific age patterns), sex, BMI, HIV infection status, household size, crowding, study site, household exposures to SARS-CoV-2, and community SARS-CoV-2 infection prevalence within the cohort. The household exposure intensity is measured as the combined shedding intensity of all actively infected household members (detailed definition in Material and Method Section 3.5). We found that the hazard of acquiring infection increased with household exposure intensity, with a stronger effect in the proliferation than the clearance phase (Figure 3C). One-unit increase in the household exposure intensity during the proliferation stage led to a 103% (95% CI 76% – 136%) increase in the hazard of infection whereas one unit increase in the clearance stage led to 58% (95% CI 41% – 77%) increase in the hazard of infection. Compared to D614G, we found that infectiousness was highest for the Delta variant, followed by the Beta variant, after adjusting for household and community exposure intensity, among other risk factors (hazard ratio against D614G: Delta 1.96, 95% CI 1.27 – 3.05, Beta 1.51, 95% CI 1.03 – 2.21). The difference between Delta and Beta’s infectiousness was not statistically different, with overlapping confidence intervals in their hazard ratios. We found that prior infection provided durable protection against reinfection throughout the study period. Compared to seronegative individuals, prior infection was 92% (95% CI 84 – 96%) protective against reinfection for the first 3 months and decreased marginally to 87% (95% CI 78 – 92%) after 9 months (Figure 3C). Individuals older than 65 years were significantly less affected during the D614G wave while children and adolescents aged 6 – 18 years were significantly more affected during the Delta wave (Figure 3C). In addition, higher BMI and residing in an urban setting were independently associated with increased risk of SARS-CoV-2 infection. We found that individuals with low CD4 cell counts (< 200) tended to be more susceptible to acquisition of SARS-CoV-2 when compared to HIV-uninfected individuals, but there was limited statistical power to detect this difference (Figure 3C). Finally, we did not find significant associations between SARS-CoV-2 infection risk and sex, household size or crowding (Figure 3C)

### Projecting the Omicron Wave and post-Omicron futures in PHIRST-C’s urban site

The study cohorts provide estimates of the duration and degree of cross-protective immunity between SARS-CoV-2 variants predating Omicron, with evidence of persistence of clinical protection beyond a year for these variants. Building on these estimates, we used mathematical models to explore a range of plausible scenarios compatible with the observed transmission dynamics of the Omicron epidemic wave in South Africa and project post-Omicron futures. Specifically, we explore how Omicron’s potential differences (relative to Delta) in infectivity, immune evasion, and severity could shape the scale and severity of the Omicron epidemic wave and the likelihood of recurrences of SARS-CoV-2 outbreaks post-Omicron. We focused the analysis on the study’s urban cohort, which is sampled from the city of Klerksdorp in Dr Kenneth Kaunda Health District located on a national transport route, and the population is well-mixed with the district population.

Details of the transmission model and calibration procedure can be found in Materials and Methods Section 4. To briefly summarize, we utilized PHIRST-C’s urban site data and district level SARS-CoV-2 surveillance information to reconstruct SARS-CoV-2’s antigen exposure(s) history in this population, considering infections and/or vaccinations through time. We then built a variant-specific transmission model to capture the declining phase of the Delta wave after the end of the PHIRST-C study and infer the population-level transmission rate during that time. Next, we added a second strain to our model to account for Omicron’s dynamics, with free parameters representing the relative infectiousness of Omicron vs Delta 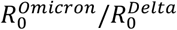, Omicron’s degree of immune evasion against infection among individuals infected by prior variants 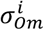, and Omicron’s degree of immune evasion against transmission given reinfection 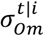 (with higher values of 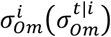 indicating higher degrees of immune evasion, where 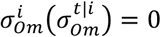 corresponds to no evasion while 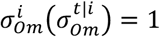 indicates 100% evasion, detailed in Materials and Methods Section 4). We explored the parameter space and initial conditions of Omicron’s introduction and selected epidemic trajectories that were compatible with the observed growth advantage of Omicron over Delta and the peak timing of the Omicron wave – the two most reliable epidemiologic observations during the Omicron epidemic. We then evaluated Omicron’s possible epidemic trajectories for the remaining parameters conditional on the observed growth rate and peak timing. Lastly, we selected a reference scenario (RS) that was most compatible with independent evidence on the degree of Omicron’s immune evasion and projected the likelihood of resurgence by different variants.

Assuming that the Delta variant was in an exponentially declining phase after week 35 of 2021 and that the Omicron variant was growing exponentially until week 48 of 2021 in the study urban district *(20)*, we estimated that the daily growth rate was -0.063 for Delta and 0.275 for Omicron, which translates into a growth advantage of 0.338 per day for Omicron over Delta (Figure S7). The transmission models of Delta and Omicron were calibrated to match their observed growth rates during this period, respectively. In Figure 4A, we show the trade-off between the estimated ratio of basic reproduction numbers between Omicron and Delta 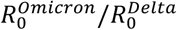 and different degrees of evasion of protection against infection 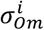 and transmission 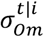. We assumed mean intrinsic generation times of 5 and 4 days for Delta and Omicron *(23)*. We found that across the full range of immune evasion parameters, Omicron had a higher basic reproduction number than Delta. However, there was clear compensation between immune evasion and intrinsic transmissibility: a higher degree of immune evasion would require a lower basic reproduction number for the Omicron variant to match the observed epidemic trajectory. We also found that for all parameters explored, the Omicron epidemic led to higher infection rates than prior epidemic waves, with the most optimistic scenario resulting in an infection rate above 40%. We project Omicron’s infection rate positively correlated with its immune evasive property (Figure 4B). Omicron infections were expected to accumulate within a relatively short period of time, with epidemic duration (measured as 4 times the standard deviation of the onset dates of all infections within the epidemic wave) projected to range from 31 – 37 days depending on the parameters. Notably, our model projected that a large fraction of Omicron infections (>40%) would be reinfections or vaccine breakthrough infections, with higher proportions observed for higher immune evasion parameters (Figure 4D). Furthermore, a low rate of clinical cases was projected for Omicron, after controlling for the number of infections due to this variant. The projected infection case ratio (ICR) for Omicron ranged from 0.4% to 0.9% (Figure 4E), much lower than the ICRs of 3.6%, 3.3%, and 9.4% estimated for the D614G, Beta, and Delta waves (Table S2). In sensitivity analyses (Figure S9), we further explored the impact of Omicron mean generation times ranging from 3-6 days *(24)*.

**Fig. 4.**
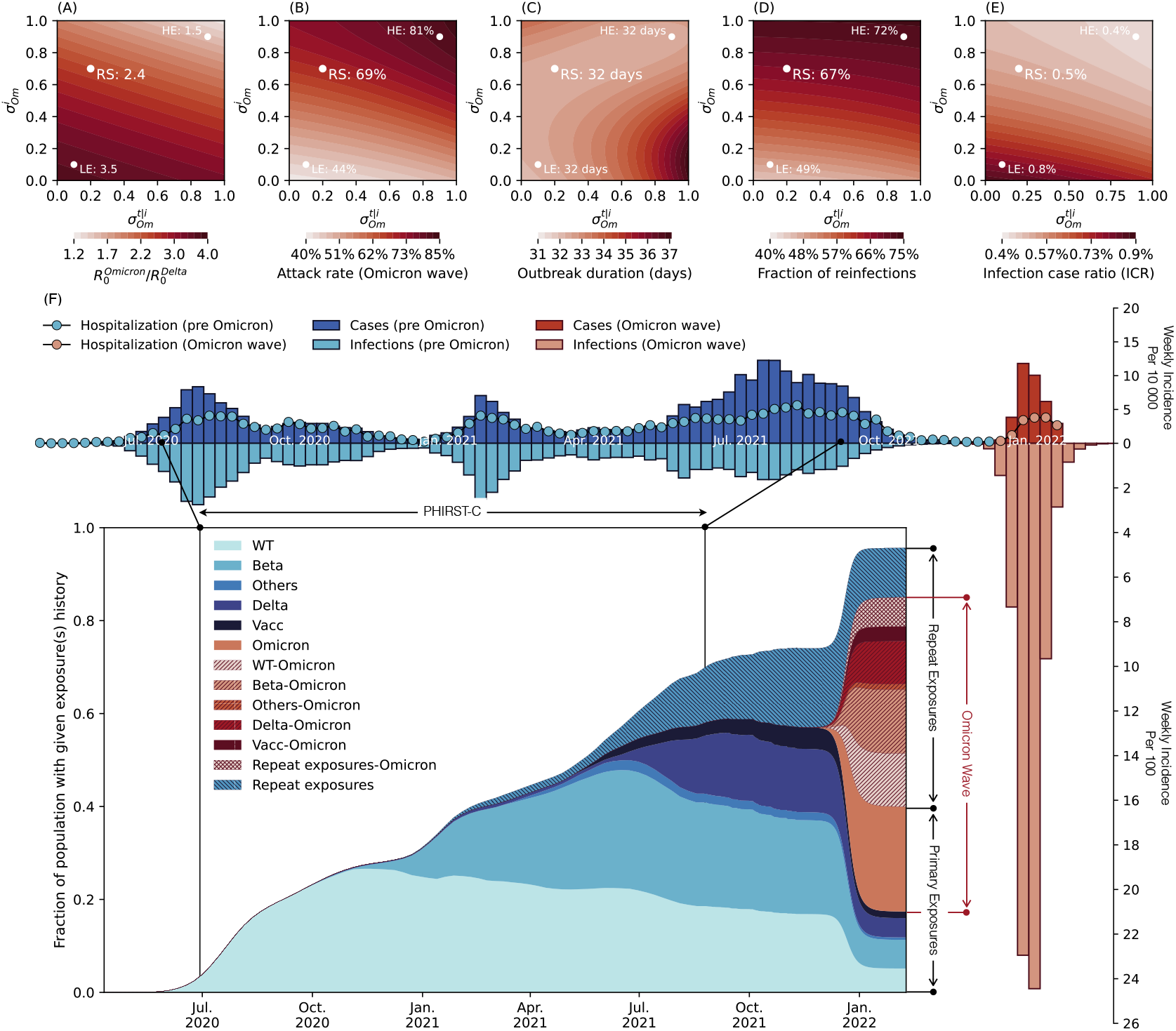
(A) Phase diagram of estimated reproduction number ratio between Omicron and Delta 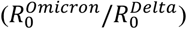 as a function of immune escape parameters 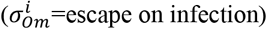 and 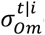 (escape on transmission reduction conditional on infection). Parameters shown are those that matched the observed growth advantage of Omicron over Delta and the timing of the Omicron peak in the urban district of the PHIRST-C study (B) Phase diagram of the infection rate of the Omicron wave as a function of 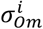 and 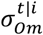 and the corresponding 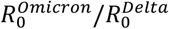 in (A). (C) Phase diagram of the epidemic duration of the Omicron wave as a function of 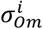 and 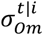 and the corresponding 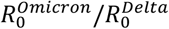 in (A). (D) Phase diagram of the fraction of reinfections of the Omicron wave as a function of 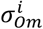 and 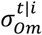 and the corresponding 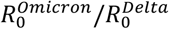in (A). (E) Phase diagram of the infection case ratio (ICR) of the Omicron wave as a function of 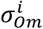 and 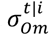 and the corresponding 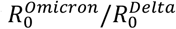 in (A). In (A)-(E), white dots marks three specific scenarios including a reference scenario (RS) with 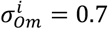 and 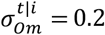, a low immune escape scenario (LE) with 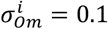 and 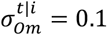, and a high immune escape scenario (HE) with 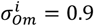 and 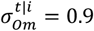. (F) For the reference scenario (RS), reconstruction of infection time series and exposure histories by variant: Top panel y axis upwards: Weekly incidence per 10,000 individuals of SARS-CoV-2 cases reported to the District till January 2022, in the period before Omicron (dark blue bars) and during Omicron (dark red bars). Top panel y axis downwards: Weekly incidence per 100 individuals of SARS-CoV-2 infections reconstructed based on PHRIST-C data (prior to September 2021) and estimated using Delta/Omicron-specific transmission models from September 2021 to the end of the Omicron wave at the end of January 2022. Pre-Omicron infections are in light blue and Omicron infections are in light red. For the top panel, the y axis upwards and downwards have different scales (by a factor of 100). Insert panel: the prevalence of the population with specific SARS-CoV-2 antigen exposure histories. Legend abbreviations: D614G: individuals who only experienced one D614G infection; Beta: individuals who only experienced one Beta infection; Delta: individuals who only experienced one Delta infection; Omicron: individuals who only experienced one Omicron infection; Others: individuals who only experienced one SARS-CoV-2 infection with genotype other than the D614G, Beta, Delta and Omicron variants; Vacc: individuals who had received at least one dose of vaccines but had not yet been infected by SARS-CoV-2; Vacc-Omicron: individuals who were vaccinated first then infected by Omicron; D614G-Omicron: individuals who were first infected by D614G then infected by Omicron; Beta-Omicron: individuals who were infected by Beta first then infected by Omicron; Delta-Omicron: individuals who were infected by Delta first then infected by Omicron; Others-Omicron: individuals who were infected by a variant other than D614G, Beta, Delta and Omicron first then infected by Omicron; Repeat exposures: individuals who were exposed to SARS-CoV-2 antigens more than once (through vaccination or infection) without Omicron infection; Repeat exposures-Omicron: individuals who were exposed to SARS-CoV-2 antigens more than twice (through vaccination or infection) then infected by Omicron.

To estimate the immune footprint of the Omicron wave and project a post-Omicron future, we considered a reference scenario (RS) for Omicron’s immune evasion characteristics, guided by independent data. We set 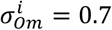 and 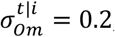, corresponding to a drop in protection against infection from 88% for pre-Omicron variants (Fig. 2J) to 47% for Omicron *(17, 25)*, and a drop in protection against transmission from 60% *(26)* to 52%, reflecting weak immune evasion on onward transmission *(18)*. Under this scenario, the estimated 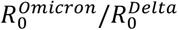 was 2.4, the infection rate was 69%, the epidemic lasted 32 days, and the fraction of reinfections and vaccine breakthroughs was 68% (Figure 4A-D, white dot). In Figure 4F, we visualize the observed incidence of reported SARS-CoV-2 cases in all four epidemic waves in the study district and report the reconstructed infection time series and population level history of SARS-CoV-2 antigen exposures. We found variable levels of under-reporting depending on the SARS-Cov-2 variant, with ICR of 3.6% (95% CI 3.4 – 3.8%) for D614G, 3.3% (95%CI 3.0 – 3.6%) for Beta, 9.4% (95%CI 8.7 – 10.2%) for Delta (Table S2), and 0.5% for Omicron’s reference scenario projection (Figure 4E). Findings for the D614G and Beta wave in agreement with previous findings *(27)*. For the reference scenario, more than 90% of the population was projected to have been infected with one or more SARS-CoV-2 variants by the end of the Omicron wave (Figure 4F). In particular, we estimated that 22.5% of the population would have seen Omicron as their first SARS-CoV-2 exposure, 16.8% would have been exposed to a pre-Omicron variant and this would remain their only SARS-CoV-2 exposure, and 45.7% would have experienced Omicron reinfections or vaccine breakthrough infections. In a sensitivity analysis, we further explored a high immune escape scenario where 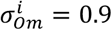 and 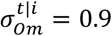 and a low immune escape scenario where 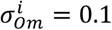 and 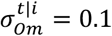. The epidemic trajectory and build-up of population immunity for the low and high immune escape scenarios are reported in Figure S5A and S5B respectively.

Because the Omicron variant is antigenically distinct from all previously circulating variants in South Africa *(14, 15, 28)*, the level of immunity conferred by Omicron’s primary and breakthrough infections/reinfections against itself, other circulating variants, and new variants, are key determinants for the likelihood of SARS-CoV-2 resurgence *(29, 30)*. To study the post-Omicron phase, we explored how different exposure histories could confer different levels of protection against infection with homologous and heterologous variants. Accordingly, in our model, the level of protection varied both with the variant that conferred immunity (either through infection or vaccine) and the hypothetical variant circulating in the post-Omicron future. Protection was expressed as a relative risk, compared to naïve individuals (where *RR*^*i*^ = 1 indicates no protection while *RR*^*i*^ = 0 indicates perfect protection). For simplicity, we assumed that protection against transmission remained constant at 60% (*RR*^*t*|*i*^ =0.4 *(26)*). We assessed the risk of future recurrences of different SARS-CoV-2 variants after the initial Omicron wave had subsided, assuming possible changes in contacts to account for erosion in adherence to SARS-CoV-2 interventions over time (models detailed in Materials and Methods Section 6).

Even at the contact rate estimated during the Delta wave, the level of population immunity would not be able to prevent a recuring Omicron epidemic unless past Omicron infection conferred high and durable protection against itself (Figure S10A). If contact rates increased 100% relative to current levels, a return of the Omicron variant would likely cause outbreaks irrespective of the protection afforded by prior Omicron infections (outbreak conditions defined as growth rate larger than zero, Figure S10B). A 100% increase in contacts may be plausible given estimated levels of transmission reduction in South Africa at the end of 2021, which reflect the combined effect of population behavior and seasonality (see Materials and Methods, Section 5.5 and Table S5). In contrast, if contact rates were to remain the same as those observed during the Delta wave, the Delta variant would be unlikely to return and cause new outbreaks, across all ranges of Omicron-specific immunity assumptions against Delta (Figure S10C). With 100% higher contact rates, some scenarios favored a re-emergence of Delta if immunity induced by Omicron did not protect well against Delta (Figure S10D). We also explored scenarios involving a hypothetical new variant X with the same basic reproduction number and generation time as the Delta variant, and at equal antigenic distance from Omicron and pre-Omicron variants. Accordingly, the relative risk of reinfection with variant X was assumed to be the same irrespective of whether an individual was primed with pre-Omicron or Omicron antigens. With an increased contact rate compared to Delta (Figure S10D), we found more opportunities for variant X to cause recuring epidemic waves in the explored ranges of parameters, primarily by escaping immunity conferred by pre-Omicron variants (Figure S10E). Emerging variants with such antigenic features need to be closely monitored in the future. On the other hand, if heterologous prime and boost infections (accounting for 45.7% of the population, Figure 4F) were found to elicit broadly protective antibodies and confer a high level of cross-protection against new variant X, only a small parameter space would be favorable to variant X epidemics (Figure S10E). Given that a large fraction of the world’s population has been primed by vaccination or infection with pre-Omicron antigens, it is important to understand how heterologous boosting by Omicron may broaden the immune repertoire, and how this could translate into clinical protection against antigenically novel variants.

## Discussion

For a period of 13 months, the PHIRST-C study carefully monitored SARS-CoV-2 infections in 222 households at a rural and an urban site in South Africa. These data provide a unique opportunity to characterize variant-specific shedding kinetics, transmission dynamics within the household, and the degree of immune protection conferred by prior infection before the Omicron surge. Longitudinal rRT-PCR data available for each infection episode at roughly 3-day resolution allowed for reconstruction of the intensity of SARS-CoV-2 exposures exerted on each household member based on *Ct* values. We found that individuals were more infectious in the RNA proliferation than clearance stage. Prior to the emergence of Omicron, substantial population immunity had accumulated through prior infection, with high and durable protection against symptomatic and asymptomatic reinfection, in line with prior findings *(31, 32)*. These detailed cohort data allowed us to project the full scope of the Omicron epidemic and assess possible futures. Overall, even with a high degree of immunity post the Omicron wave (with over 90% of the population previously exposed to SARS-CoV-2 antigens), recurrence of past or antigenically novel variants is plausible, especially if post-Omicron behavioral changes increase contacts.

The disease burden of future SARS-CoV-2 epidemic waves depends on the intrinsic severity of the variants themselves as well as the level of protection conferred by pre-existing immunity *(33)*. Current evidence suggests that while Omicron is able to evade immunity against infection to a significant degree, protection against severe outcomes remains high *(17, 34, 35)*. Our model’s projections also suggest a low clinical burden of Omicron, with a greater than 10-fold reduction in infection case ratio relative to prior waves (Figure 4E, Table S2). If existing immunity can sustain protection against severe outcomes over long timescales, the disease burden of future epidemic waves would be attenuated even if infections were widespread. However, if protection against severe outcomes waned over time, vaccine boosting would likely be needed to compensate for loss of protection.

As the PHIRST-C sampling scheme was not symptom-driven, it allowed us to capture shedding kinetics in both symptomatic and asymptomatic individuals. We observed that most infections 86.8% (581/669) were asymptomatic, but asymptomatically infected individuals transmitted the virus within their households. For context, in the South African winter seasons of 2017-2018, approximately half of the influenza infections within the PHIRST-C cohorts were asymptomatic *(36)*. The transmission potential of asymptomatic or pre-symptomatic SARS-CoV-2 infections identified in our study is in sharp contrast with SARS-CoV-1, where most transmission occurs after symptom onset *(37)*. It is also worth noting in our cohort data, prior infections, whether symptomatic or not, conferred durable protection against reinfection. We found that the shedding kinetics of SARS-CoV-2 were characterized by a rapid RNA proliferation stage until peak viral load, followed by a more gradual RNA clearance stage. The median duration of rRT-PCR positivity lasted 10.5 days (IQR 6.3 – 14.0 days) with median peak Ct = 23.1 (IQR 20.0 – 27.2), in agreement with other high-frequency sampling studies *(21, 22)*. Reinfections had shorter durations of rRT-PCR positivity and lower shedding peaks compared to primary infections (Figure 2 H-I), which would be expected to decrease the probability of onward transmission. Our findings align with reports of reduction in viral shedding among vaccine breakthroughs relative to primary infections, prior to the occurrence of the Omicron variant *(38–41)*. Interestingly, we observed variation in infectiousness through the course of infection, after adjusting for Ct values, whereby an individual in the proliferation stage tends to be more infectious than one in the clearance stage. The post-peak decline in infectiousness coincides with the onset of adaptive immune responses that work to suppress the on-going infection *(42)*. The observed decline in infectiousness in the RNA clearance stage also could be due to neutralization of some viral particles by antibodies, precluding productive transmission.

The peak of COVID-19 hospitalizations during the Omicron wave was lower than that during the Delta wave in South Africa *(43)*, despite our model projecting a much higher infection rate for Omicron than Delta. This is compatible, however, with some lines of evidence suggesting a lower severity of the Omicron variant in naïve individuals, combined with robust infection and vaccine-induced immunity against severe Omicron disease *(17, 44)*. However, it is important to stress that, in South Africa, immunity accumulated prior to the Omicron wave was mostly through prior infections, due to a delayed start and slow rollout of the vaccine campaign *(45)*. As a result, the proportion of the population infected by pre-Omicron variants was substantially higher in South Africa than in countries that experienced faster vaccine rollout and/or effective mitigation strategies.

Our study have several limitations. First, our findings about the persistence of infection-induced immunity are based on a 13-month study. The duration and quality of protective immunity over longer timescales remain open questions. Recent studies have found that antibody responses improve over time through affinity maturation *(46, 47)* and that long-lived plasma cells can be identified in the bone marrow at least one year after infection, suggesting that immunity conferred by infection or vaccination could be potent and durable against non-immune evasive variants *(48)*. Persistent germinal center responses and durable T cell memory have also been observed among vaccine recipients *(49–51)*. However, how the protection holds up against immune-evasive variant such as Omicron remains an outstanding question. Unfortunately, the PHIRST-C cohorts did not cover the Omicron wave, thus we could not directly measure immune protection at the individual level and relied on modelling of population-level dynamics. Post-Omicron serologic surveys following up the cohort population could provide deeper insight into the full impact of the Omicron wave. In our projections of SARS-CoV-2 resurgences, we did not consider waning explicitly since our cohort data did not support pronounced waning of infection-induced immunity. Accordingly, our projections are most relevant to short time scales, in the order of a few months. Interestingly, we find that resurgences are likely even over short time horizons. A second limitation relates to missed infection episodes, despite frequent rRT-PCR testing. In total, 21% (303407 person-days/1472400 person days) of the total person-days of observation were excluded from the regression during the entire study period due to missing nasal swab visits, missing serologic status, or experiencing an active infection episode. For example, 14% (90/639) of individuals who seroconverted during the study lacked rRT-PCR confirmation of active infections (i.e., their first serologic test was negative, but they seroconverted later). This could possibly be due to 1) delayed seroconversion from infection episodes occurring prior to the first nasal specimen, 2) infection episodes that occurred during missed routine household visits, 3) a shorter duration of rRT-PCR positivity than the interval between consecutive nasal swabbing (3 days), 4) false positives in serology test results or 5) false negatives of the rRT-PCR assay specimen (due to specimen quality issues or detection limit of the rRT-PCR). A third limitation relates to the simplicity of the contact structure in our transmission models. Projections of the trajectories of Delta, Omicron, and variant X did not address heterogeneity due to age-specific susceptibility, transmissibility, and contact patterns. Nor did we consider individual variation in infection and vaccine-derived protection. Heterogeneity in mixing patterns and immune protection could lead to a lower infection rate when compared to homogeneous models with the same basic reproduction number *(52, 53)*, thus the size of the Omicron epidemic could be inflated in our projections. In addition, our size projections could be inflated if the Omicron’s serial interval was shorter (corresponding to a lower basic reproduction number) than that the range of values explored. Population surveys on active infections during the Omicron wave as well as paired-sera surveys before and after the Omicron wave will be necessary to confirm the true scale of the Omicron epidemic.

In conclusion, our study provides an in-depth analysis of the kinetics of viral shedding, transmission dynamics, and persistence of immunity conferred by sequential exposures to different SARS-CoV-2 variants, and how these factors contribute to shaping the Omicron and post-Omicron phases. We found durable cross-protective immunity conferred by prior infection against pre-Omicron variants. However, Omicron successfully breached population immunity due to a combination immune escape and increased transmissibility, reinfecting a large fraction of the population and leaving a complex immune landscape in its aftermath. With increasing contacts as the Omicron wave subsides, several possible scenarios for SARS-CoV-2 recurrences are possible, involving both old and new variants. Further work on how immunity may strengthen and broaden upon sequential exposures with different variants and vaccination episodes will be important to clarify the next phase of the pandemic.

## Data Availability

The investigators welcome enquiries about possible collaborations and requests for access to the data set. Data will be shared after approval of a proposal and with a signed data access agreement. Investigators interested in more details about this study, or in accessing these resources, should contact the principle investigator, Prof Cheryl Cohen, at NICD.

## Acknowledgment

The findings and conclusions in this report are those of the authors and do not necessarily represent the official position of the NIH or the U.S. Centers for Disease Control and Prevention.

## Author contributions

KS, ST, JK, AvG, MLM, NW, JM, NAM, KK, STo, LL, CV, CC designed the experiments. CC, JK, and ST accessed and verified the underlying data. ST, JK, AvG, MLM, NW, JNB, JM, MdP, MC, AB, NAM, KK, STo, LL, FW, JdT, FXG, FSD, TMK, CC collected the data and performed laboratory experiments. KS, ST, JK, AvG, MLM, NW, JNB, JM, MdP, MC, AB, NAM, KK, STo, LL, FW, JdT, FXG, FSD, TMK, CV, and CC analyzed the data and interpreted the results. KS, ST, JK, AvG, CV, and CC drafted the manuscript. All authors critically reviewed the Article. All authors had access to all the data reported in the study.

## Ethics statement

The PHIRST-C protocol was approved by the University of Witwatersrand Human Research Ethics Committee (Reference 150808) and the U.S. Centers for Disease Control and Prevention’s Institutional Review Board relied on the local review (#6840). The protocol was registered on clinicaltrials.gov on 6 August 2015 and updated on 30 December 2020 (https://clinicaltrials.gov/ct2/show/NCT02519803). Participants receive grocery store vouchers of ZAR50 (USD 3) per visit to compensate for time required for specimen collection and interview.

### Supplementary Materials

Materials and Methods

Figures S1-S10

Tables S1-S7

## Supplementary Materials

### Materials and Methods

#### 1. Cohort design

We conducted a prospective household cohort study of SARS-CoV-2 transmission at an urban and a rural site in South Africa from July 2020 to August 2021*(4)*. The rural site was in Agincourt, a rural community in Mpumalanga Province, which has been a longstanding health and socio-demographic surveillance system site. The urban site was in Klerksdorp, an urban community located in the North West Province. This study was built upon the larger multi-year **P**rospective **H**ousehold cohort study of **I**nfluenza, **R**espiratory **S**yncytial virus and other respiratory pathogens community burden and **T**ransmission dynamics (PHIRST), which was conducted from 2016 to 2018 to monitor transmission of respiratory pathogens *(54)*. The study was repurposed for SARS-CoV-2 during the pandemic. To study infection and reinfection with SARS-CoV-2, a total of 222 households (114 in the rural site and 108 in the urban site) with at least 3 household members were enrolled between July 2020 and August 2021, consisting of 638 and 557 participants in the rural and urban site, respectively. In the rural site, we first approached households from the 2017 and 2018 cohorts, and in the urban site, those from the 2016, 2017 and 2018 cohorts. To supplement the sample size of the cohorts, we enrolled additional households at each site using the same methods as for the initial PHIRST study. Baseline demographic factors and information on underlying medical conditions were collected at enrollment (Table 1). Throughout the study period, household members were visited twice a week by study nurses and trained lay field workers for collection of biological and clinical data. At each visit, upper respiratory tract specimens were collected using mid-turbinate nasal swabs, irrespective of symptom presentation. Data on symptoms, healthcare seeking behavior, hospitalization and death were captured at each follow up visit on a REDCap tablet-based real-time database. Respiratory specimens were tested by rRT-PCR for SARS-CoV-2. The lineage types of positive rRT-PCR specimens were determined by variant-specific rRT-PCR assay. Sera were collected at enrollment and approximately every 2 months during the 11-month follow-up period from all participants (see Figure 1 for timeline) and tested for the presence of SARS-CoV-2 antibodies.

#### 2. Laboratory methods

The detailed laboratory methods were previously described *(4)*. To briefly summarize, nucleic acids for real-time reverse transcription polymerase chain reaction (rRT-PCR) tests were extracted using the Hamilton Microlab NIMBUS Instrument (Hamilton, Nevada, USA) with the STARMag Universal Cartridge kit (Hamilton, Nevada, USA) and the STARMag Universal Cartridge kit (Seegene Inc., Seoul, Korea) according to the manufacturer’s instructions. Specimens were tested for the presence of SARS-CoV-2 by rRT-PCR using the Seegene Allplex™ 2019-nCoV kit (Seegene Inc., Seoul, Korea). Initial positive specimens were re-extracted and tested again. Only specimens with at least two out of three gene targets confirmed positive during the second test were considered as positive specimens. The cycle threshold (Ct) value of each rRT-PCR test was recorded for further analysis. SARS-CoV-2 lineage was determined through Seegene variant I and II typing assays which differentiate variants Alpha (B.1.1.7), Beta (B.1.351), Delta (B1.617.2) and Gamma (P.1) *(4)*.

Serum specimens were collected using venous blood, centrifuged into serum separator tubes and stored refrigerated immediately and transported to NICD. Aliquots of prespecified volume according to manufacturer instructions were tested for the presence of SARS-CoV-2 antibodies by the Roche Elecsys Anti-SARS-CoV-2 assay against nucleocapsid (N) antigen *(55)*. Assay readout above or equal to cutoff index 1 is considered as sero-positive, while below cutoff index 1 is considered as sero-negative. Negative control validations were performed using serum specimens from participants at both study sites prior to 2020 *(4)*. An independent study benchmarking performances of available commercial and laboratory serologic assays demonstrated that the Roche Elecsys Anti-SARS-CoV-2 assay had high sensitivity and specificity across a wide range of severity spectrum for at least 6 months post-infection *(56)*.

#### 3. Statistical analysis

##### 3.1 Seroconversion and vaccination

Combining the longitudinal rRT-PCR assays and serological test results from the two study sites, we found that the seroconversion rate was 97.6% (583/600) among primary infection episodes with at least one blood specimen collected 30 days after their first rRT-PCR positive test. Among 639 individuals with negative serological specimen(s) after their first blood specimen was taken but who seroconverted later, 86% (549/639) were confirmed by rRT-PCR during the study period, suggesting that twice-weekly rRT-PCR testing captured most of the infections during the study period. Among 706 seroconverted individuals with at least one follow-up blood specimens after seroconversion, only 40 (5.7%) later sero-reverted (sero-reversion is defined as sero-converted individual who became sero-negative the subsequent serologic test). The baseline characteristics of all individuals are reported in Table 1. 10% individuals received at least one dose of SARS-CoV-2 vaccine and 5% were fully vaccinated within the study period of PHIRST-C.

##### 3.2 Defining and typing variant-specific infection episodes

For each individual in the study, we defined the duration of a SARS-CoV-2 infection episode as the time interval between the first and the last of a set of positive rRT-PCR test, where consecutive positive tests were separated by less than 30 days. If at least one positive rRT-PCR test within the infection episodes was identified as any of the variants of concern (VOC) (defined by the WHO) Alpha (B.1.1.7), Beta (B.1.351), Delta (B.1.617.2), the lineage of the infection episode was assigned to the identified VOC. If all lineage-typed rRT-PCR tests within an infection episode were identified as the D614G, then the lineage of the infection episode was assigned to D614G. If none of the positive rRT-PCR specimens within an infection episode had a lineage defined, the lineage of the infection episode was labeled as inconclusive. In total, we observed 669 infection episodes, including 634/669 (95%) with defined lineages. For the 35/669 (5%) infection episodes with inconclusive lineages, we assigned the infection to the dominant SARS-CoV-2 lineage at the study site identified within the month of the infection episode. After lineage extrapolation, 108/669 (16%) of the infection episodes were estimated to be caused by D614G lineage; 245/669 (37%) infection episodes were estimated to be caused by VOC Beta lineage; 299/669 (45%) infection episodes were estimated to be caused by VOC Delta lineage; 17/669 (3%) were estimated to be caused by other lineages.

##### 3.3 Characterizing the relative viral RNA shedding kinetics of SARS-CoV-2 infection episodes

When interpreting Ct values from rRT-PCR test by the Seegene Allplex™ 2019-nCoV kit, it is important to stress that the Ct value of a single rRT-PCR test is not a direct measurement of the quantity of viral genetic material present in an individual specimen (in absolute terms) *(57)*. Many factors could influence the Ct value of a rRT-PCR test, including but not limited to the specimen quality, extraction method, chemistry of reagents, gene targets. Further, Ct values cannot be directly compared between assays of different types *(58)*. However, comparing serial Ct values and/or Ct values of different groups of population collectively, based on rRT-PCR tests from the same assay and laboratory setting, does reflect the relative variation in terms of viral genetic material concentration over time and between population subgroups *(57, 58)*.

We measure the viral RNA shedding intensity of a given specimen as the cycle threshold value of Seegene Allplex™ 2019-nCoV kit’s N gene target Ct value of the specimen, i.e., 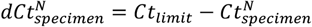. For the Ct value, we are only considering the N gene target to avoid between-target variation. In this study, instead of directly interpreting the Ct value of a single rRT-PCR test, we fit a mathematical model to capture the temporal kinetics of *Ct* for longitudinally collected specimens and extract statistical summaries of RNA shedding intensity for each infection episode. In particular, following the method presented in *(21)*, for each infection episode, we modelled the *Ct* kinetics during the “RNA proliferation stage”, characterized by linear decrease in *Ct* until a trough in *Ct* is reached; and during the “RNA clearance stage”, characterized by linear increase (in *Ct*) from the trough until the last rRT-PCR positive result. The duration of the “RNA proliferation stage” *τ*_*p*_ was defined as the time from *Ct* first exceeding the detection threshold (i.e.,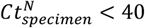) until the trough of *Ct*; the duration of the “RNA clearance stage” *τ*_*c*_ was defined as the time from trough *Ct* to Ct reaching above detection threshold (i.e.,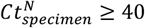). The duration of rRT-PCR positivity *τ*_*s*_ for each infection episode was defined as the total duration of the RNA proliferation and RNA clearance stages: *τ*_*s*_ = *τ*_*p*_ + *τ*_*c*_. The duration of RNA proliferation *τ*_*p*_, RNA clearance *τ*_*c*_, rRT-PCR positivity *τ*_*s*_, as well as the trough shedding intensity 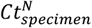 were estimated for each infection episodes using Markov Chain Monte Carlo (MCMC) method.

The fit of the RNA shedding trajectories for each variant are shown in Figure 2A-G. In addition, we performed multivariable regression analysis to evaluate the dependencies of duration of rRT-PCR positivity and trough *Ct* on participants’ characteristics including age, sex, BMI, HIV infection status, symptom status, variant type, and evidence of prior infection. Since the nasal swab sampling period ended on August 28, 2021, around the peak of the Delta wave in both sites, we limit our analysis to infection episodes with first positive PCR sample 30 days prior to the end of sampling to avoid censoring bias. The result of the regression is presented in Figure 3A-B.

##### 3.4 Assigning the lineage of prior infections among seropositive individuals

For individuals who seroconverted without a rRT-PCR confirmed infection episode (prior to the start of the PHIRST-C cohort), we assigned the lineage type of the individual’s unobserved infection according to lineages’ prevalence (based on the infection episodes that had lineage information) at the study site in the month of the earliest seropositive specimen.

##### 3.5 Risk factors associated with SARS-CoV-2 (re)infections

For each individual in the urban and rural sites, we first reconstructed the status of prior and ongoing infections at a daily time resolution, based on the lineage-typed infection episodes and serologic results described in Section 3.3 and 3.4 above. Specifically, we denoted a person-day observation of individual *i*’s infection status at day *t* as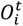. We set 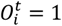 if individual *i* acquired infection on day *t* (i.e., the date of viral acquisition, marking the start of an infection episode), and set 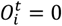 if individual *i* didn’t acquire infection on day *t*. Individuals without rRT-PCR positive results throughout the study period were assumed to escape infection throughout the observation period (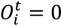, for all t). For individuals with observed infection episodes during the study period, the individual Ct kinetics were modelled as in Section 3.3. The model allowed us to estimate the time at which Ct crossed the rRT-PCR detection limit (*Ct* = 40). We assumed that the time *t*_*inf*_ of acquiring infection 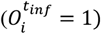occurred *δT*_*cryptic*_ = 4 days prior to the Ct crossing the detection threshold *(59)*. The time period from infection onset to 30 days after viral RNA clearance (i.e., the estimated time of Ct crossing below the rRT-PCR detection limit at cycle threshold of 40) was considered a period of active infection, when an individual cannot get infected again. Thus, this refractory period was excluded from the survival analysis of (re)infection hazard. We also considered a sensitivity analysis with a 15-day exclusion after viral RNA clearance. We also censored participants after they received their first dose of vaccine. In Figure S1, we visualize the vaccine uptake overtime for both J&J/Janssen Ad26.COV2.S and the Pfizer/BioNTech BNT162b2.

After having reconstructed the daily infection status of each individual, we modeled the risk of (re)infection for each individual using piecewise exponential models of survival analysis with time varying covariates *(60, 61)*. The piecewise exponential model is estimated by performing Poisson regression with the daily infection status of each individual as the binary outcome and daily covariates *(60, 61)*. The covariates considered in the regression analysis included the individual’s age (allowing for variant-specific age effects), sex, body mass index (BMI), HIV infection status, household size, household crowding, variant type, study site, time since prior infection, SARS-CoV-2 exposure intensity from household members with on-going SARS-CoV-2 infection, and SARS-CoV-2 prevalence in the community. We estimated that the SARS-CoV-2 exposure intensity from household members with on-going SARS-CoV-2 infections at time *t* as the sum of the estimated *dCt*s of all infected household members *(62)*. The *dCt* of household member *j* at time *t* was measured as 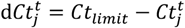. Thus, the household exposure intensity 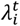 exerted on individual *i* at time *t* can be expressed as 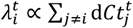, where *j* is the sum over all infected household members at time *t*. We further discriminated the household exposure intensity during the RNA proliferation and RNA clearance phases of infections in household members. We estimated the weekly SARS-CoV-2 community prevalence as the prevalence of each variant in the entire study site (no of variant-specific infections that week/population tested that week) and use the variant-specific prevalence as a proxy for the SARS-CoV-2 community exposure intensity. We included household- and individual-level hierarchical random effects, as well as “day-of-week” and “day-of-year” random effects in the regression model. The full model selection procedure is reported in Table S1 (the model selected was model 16, corresponding to regression results presented in Figure 3C).

We considered whether saturation effects could possibly affect our estimates of infection risk in the proliferation and clearance phases. If the risk of transmission in the household setting was very high during the proliferation stage, so that most household members were infected at the end the proliferation stage of the index case, there may not be enough observations to power the estimate of transmission hazard during the clearance stage. Empirically, however, we found that transmission saturation was uncommon for the participating households. We define a household transmission cluster as a group of infections separated no more than 14 days by their time of infections (equivalent to hierarchical clustering based on infected individuals’ time of infection with single-linkage). In Figure S3, we plot the distribution of the size of household clusters as a function of the number of individuals in the household (household size). We found than only 10 household transmission clusters were saturated (all members were infected), and the median size of the household cluster was 38% (proportion of household members infected). Thus, saturation was not an issue, and there were enough susceptible individuals to power the estimation of the hazard of transmission form infected individual during the clearance phase of infection.

Since the nasal swab sampling period ended on August 28, 2021, around the peak of the Delta wave in both sites, we limit our analysis to infection episodes with first positive PCR sample 30 days prior to the end of sampling to avoid censoring bias. In total, 21% (303407 person-days/1472400 person days) of the total person-days of observation were excluded from the regression during the entire study period due to missing nasal swab visits, missing serologic status, or individuals experiencing an active infection episode. A household with an individual with chronical SARS-CoV-2 infection was also excluded. The hazard ration of the piecewise exponential model is presented in Figure 3C (corresponding to Model 16 in Table S1). We conducted two additional sensitive analyses with slight variation of this model: in the first sensitivity analysis (Figure S4A) we censored individuals until 15 days post viral RNA clearance (instead of 30 days in the main analysis); in the second one (Figure S4B) we used logistic regression (logic link) rather than Poisson regression (log-link) *(63)*.

**Table S1.**
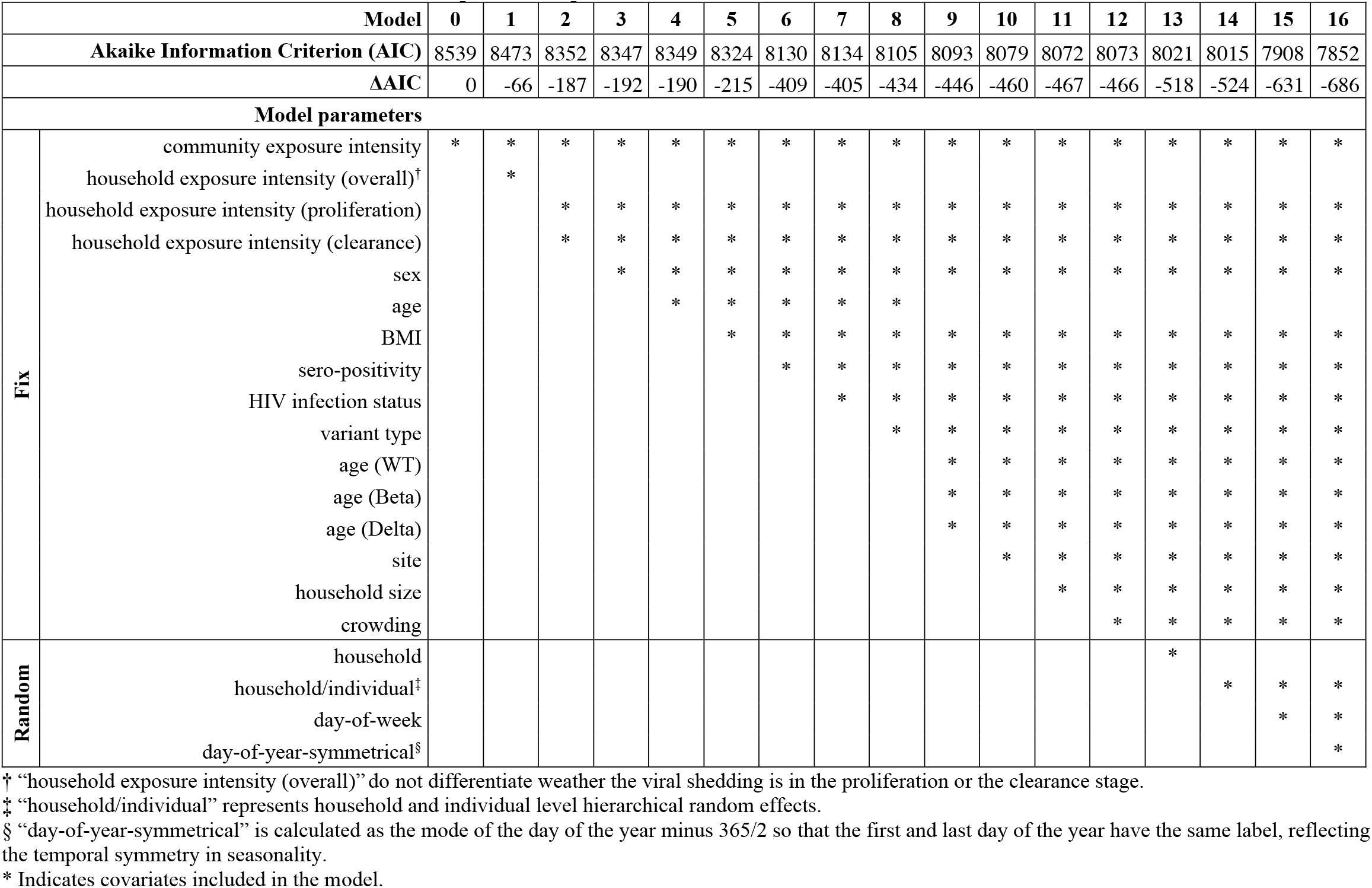
Model selections on the covariates of the piecewise exponential model for the individual risk of (re)infection.

#### 4. Reconstructing the SARS-CoV-2 prior infection/vaccination history in the Dr Kenneth Kaunda District by September 2021

In this Section, we use PHIRST-C’s urban site as a sentinel for SARS-CoV-2 antigen exposure(s) history (including both infection and vaccination) for the broader district where the study is located (Dr Kenneth Kaunda District). We reconstruct the population-level prevalence of different type of antigen exposures over time before Omicron and use this information as the initial condition in models that project the impact of Omicron. We focus on the urban site (Klerksdorp) as it is a major city of the Dr Kenneth Kaunda District.

##### 4.1 Estimating the reporting rates of D614G, Beta, and Delta during the first three epidemic waves in the Dr Kenneth Kaunda District

Here we compare the carefully monitored weekly PHIRST-C infection data with weekly surveillance data for the broader district to estimate aspects of SARS-CoV-2 dynamics in the district, including the rate of under-reporting for each of the pre-Omicron variants, pre-Omicron infection histories, and the impact of the Omicron wave. The dark blue bars in Figure 4F top panel shows the weekly incidence *I*_*case*_(*t*) of SARS-CoV-2 cases in the Dr Kenneth Kaunda District, North West Province, South Africa during the first three epidemic waves captured by SARS-CoV-2 surveillance. We calculate the weekly cumulative case rate *C*_*case*_(*t*) at time *t* as 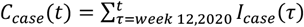, where week 12, 20202 is the week when the district starts reporting SARS-CoV-2 cases. We denote the cumulative infection attack rate at time *t* (proportion of the population infected by each of these variants at time t) for D614G, Beta, Delta (and others) variant as 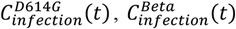, and 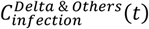 respectively. We estimate 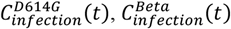, and 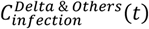 within the time period of the PHIRST-C cohort based on PHIRST-C’s serology and variant-typed SARS-CoV-2 infection episodes (Detailed in Materials and Methods Section 3). We denote the reporting rate of D614G, Beta, and Delta (equal to Others) in the urban site as *β*_*D*614*G*_, *β*_*Beta*_, and *β*_*Delta & Others*_. We assume that the reporting rate for each variant (within each epidemic wave) is constant and that Other variants (low frequency, mostly in between second and the third waves. See Figure 1F) have the same reporting rate as Delta. The cumulative case rate *C*_*case*_(*t*) and cumulative infection rates 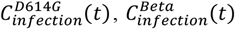, and 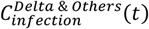 at time *t* satisfy 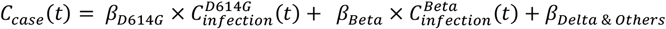. Given 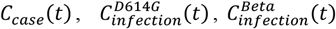, and 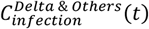 for *t* within the PHIRST-C cohort, we can estimate *β*_*D*614*G*_, *β*_*Beta*_ and *β*_*Delta & Others*_ using linear regression without intercept. The regression analysis was performed using R package lme4 version 1.1-27.1 *(64)*.The estimates are reported in Table S2.

**Table S2:** Reporting rate for each variant during the first three epidemic waves in Dr Kenneth Kaunda district, estimated by comparing infections in the PHIRST study with passive surveillance data in the broader district.

| Parameter | Estimate | 95% CI |
| --- | --- | --- |
| $\beta_{D614G}$ | 0.036 | 0.035 – 0.038 |
| $\beta_{Beta}$ | 0.033 | 0.030 – 0.036 |
| $\beta_{Delta \& others}$ | 0.094 | 0.087 – 0.010 |

##### 4.2 Estimating the weekly incidence rate of SARS-CoV-2 infection based on case incidence and reporting rate in Dr Kenneth Kaunda

Based on the variant-specific reporting rate *β*_*X*_ (*X* ∈ {*D*614*G, Beta, Delta & Others*}) estimated in Section 4.1 and the variants’ proportions *p*_*x*_(*t*) at a given time point *t*, we can express the overall reporting rate *β*_*Overall*_ (*t*) at time *t* as:

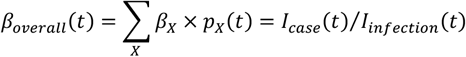

Where *I*_*case*_(*t*) is the weekly incidence rate of SARS-CoV-2 cases reported to the Dr Kenneth Kaunda District at time *t* and *I*_*infection*_ (*t*) is the weekly incidence rate of SARS-CoV-2 infections. We can thus estimate *I*_*infection*_ (*t*) as

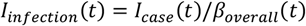

The estimated *I*_*infection*_ (*t*) prior to the end of PHIRST-C (September 2021) is visualized in Figure 4F top panel (light blue bars prior to September 2021).

##### 4.3 Reconstructing the SARS-CoV-2 antigen exposure history in Dr Kenneth Kaunda by September 2021

In addition to allowing to trace SARS-CoV-2 infection history in detail, the PHIRST-C cohort also recorded participants’ timing of vaccinations. Specifically, at time *t*, we denote the proportion of the PHIRST-C urban site population with a single prior SARS-CoV-2 infection, which is to D614G, as 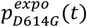 with 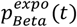 and 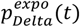 representing the same quantities for Beta and Delta. We denote past vaccination (at least one dose) as 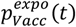 with repeat exposures (including repeat infections, vaccination followed by infection, or infection followed by vaccination) as 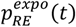. Then we can express the proportion of population with past SARS-CoV-2 infections at time t as:

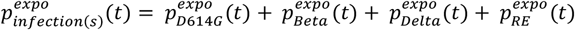

If we denote the cumulative infection attack rate in Dr Kenneth Kaunda 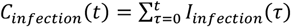 and assume the PHIRST-C urban site is a representative survey of the population in Dr Kenneth Kaunda, we can express the cumulative prevalence of past exposure of a given exposure type *X* at time *t* as:

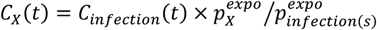

Where *X* ∈ { *D614G, Beta, Delta, Repeat Exposures, Vaccinated* }. In Figure S6 (below) and Figure 4F bottom panel, we visualize the proportion of population with a specific SARS-CoV-2 antigen exposure history.

#### 5. Modelling the transmission dynamics of the Delta and Omicron variant from September 2021 to the end of Omicron wave

##### 5.1 Overview of protective immunity

To project the impact of Omicron, we need to model how immunity from infection with pre-Omicron variants and/or vaccination will impact the probabilities of infection, transmission and severe outcomes. The effectiveness of protective immunity (*IE*) can be measured very broadly as *IE* = 1 − *RR* where *RR* is the relative risk of an outcome of interest (infection, transmission, hospitalization, etc.) and the comparison is against individuals with no prior immunity or with different types of immunity. In particular, we consider three aspects of protection from prior infection/vaccination:

1. Prior infection/vaccination could reduce the host’s susceptibility to reinfection *IE*^*i*^, measured as *IE*^*i*^ = 1 − *RR*^*i*^ where *RR*^*i*^ is the relative risk of reinfection/breakthrough infection when compared to the infection risk in a naïve population, controlling for same level of exposure.
2. Prior infection/vaccination could reduce the risk of onward transmission given reinfection/breakthrough infection *IE*^*t* |*i*^, measured as *IE*^*t* |*i*^ = 1 − *RR*^*t* |*i*^ where *RR*^*t* |*i*^ is the relative risk of onward transmission for reinfections/breakthrough infection when compared to that of primary infections, conditional on the same contact rate. Here *RR* ^*t* |*i*^ = *RR* ^*td*|*i*^ × *R* ^*ts*|*i*^ can be further broken down into the product of reduction in the duration and intensity of shedding (*RR* ^*td*|*i*^ and *RR* ^*ts*|*i*^).
3. Prior infection/vaccination could reduce the risk of disease given reinfection/breakthrough infection. The concept of COVID-19 disease is generic and could encompass a wide spectrum of severity endpoint including symptomatic cases, hospitalizations, and deaths. In this study, we used symptomatic illness as the severity end point. The effectiveness of protective immunity against being symptomatic case, conditional on infection can be measured as *IE*^*c* |*i*^ = 1 − *RR* ^*c* |*i*^, where *RR* ^*c* |*i*^ is the relative risk of disease for reinfections/breakthrough infections compared to primary infections.

##### 5.2 Modelling boosting and evasion of protective immunity in the Omicron era

Repeat exposures to SARS-CoV-2 antigens can result from repeated infections, booster shots following primary vaccine schedule(s), infection following vaccination or vice versa. Multiple exposures could stimulate a recall response and boost the level of protective immune effectiveness *IE* through reducing *RR* further from the baseline protective immunity provided by primary exposure. For example, let’s denote the protective immunity conferred by primary infection with a pre-Omicron (pOm) strain as *IE*_*pOm*_ (*pOm*) = 1 − *RR* _*pOm*_ (*pOm*). We can express the level of protective immunity conferred by reinfection with pre-Omicron strains as 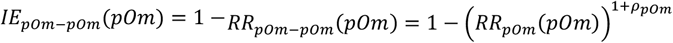, with *ρ*_*pOm*_ > 0 measuring the degree of immunity boosted by reinfection with a pre-Omicron strain. For *RR*_*pOm*_ (*pOm*) < 1), the larger *ρ*_*pOm*_, the greater the reduction *RR*_*pOm*_ (*pOm*),when compared to *RR*_*pOm*_(*pOm*), and the higher the protection conferred by boosting.

On the other hand, in face of an antigenically distinct variant like Omicron, prior immunity may not be as efficacious due to reduced ability to recognize the antigen through immune memory, leading to elevation in *RR* and consequently reduction in *IE*. For example if we denote the protective immunity conferred by primary infection with a pre-Omicron strain against refection by pre-Omicron as 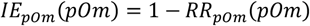, we can express the level of protective immunity conferred by pre-Omicron strain infection against Omicron (Om) as 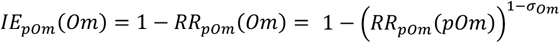, where *σ*_*Om*_ measures the degree of immune evasion by Omicron. When *σ*_*Om*_ = *IE*_*pOm*_(*Om*) = *IE*_*pOm*_(*pOm*), indicating no immune escape. When *σ*_*Om*_ = 1, *IE*_*pOm*_(*Om*) = 0, indicating 100% immune escape. We can further model the combined effects of boosting and immune escape. For example, if we consider an individual first infected with a pre-Omicron strain, followed by an Omicron infection, we can express the individual’s level of protective immunity against further Omicron infection as 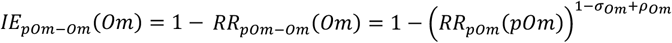.

##### 5.3 Notation conventions and an exhaustive list of different types of protective immunity considered

Following Section 5.1 and 5.2, we use 1 − *RR* as a measurement of the level of protective immunity. In particular, 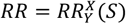 can be broken down into three independent dimensions: 1) the type of protective immunity - (superscript), 2) the antigen exposure history *Y* that confers immunity (subscript), 3) the viral strain *X* against which immunity is directed (brackets).

Where *X* ∈ {*i, t*|*i, td*|*i, ts* |*i, h*|*i*} and:

- *i* denotes protection in terms of susceptibility against infection.
- *t*|*i* denotes protection against transmission given infection.
- *td*|*I* denotes protection against transmission through reduced duration of shedding, given infection.
- *h*|*i* denotes protection against transmission through reduced intensity of shedding, given infection.
- *h*|*i* denotes protection against hospital admission given infection.

Where *Y* ∈ {*pOm, Om, pOm* − *pOm*, − *Om* } and:

- *pOm* denotes immune histories including only one antigen exposure by either pre-Omicron (including D614G, Alpha, Beta, Delta, and other non-Omicron variants in South Africa) infection or a primary schedule of vaccination.
- *Om* denotes immune histories consisting in a single exposure to SARS-CoV-2 via an Omicron infection.
- *pOm* − *pOm* denotes immune histories including at least two antigen exposures with first exposure being either pre-Omicron strain or vaccination and second exposure either pre-Omicron strain or vaccination.
- *pOm* − *Om* denotes immune history including at least two antigen exposures with the first exposure being either pre-Omicron strain or vaccination and the second exposure being an Omicron infection.

Where *S* ∈ {*pOm, Om*} and:

- *pOm* denotes protection against pre-Omicron strains.
- *Om* denotes protection against Omicron.

In this study, since the antigenic difference between Omicron and all previously circulating strains (D614G, Alpha, Beta, Delta) is much larger than antigenic differences among previously circulating strains *(17, 25, 31)*, we do not differentiate immunity between pre-Omicron variants (all pre-Omicron variants confer the same type of homologous and heterologous immunity to each other, and we also consider the vaccine to be antigenically similar to pre-Omicron strains since it is based on wild type strain). For simplicity, considering the low vaccination rate in South Africa, we assume that a full vaccination schedule confers similar levels of protection as an infection with a pre-Omicron strain (i.e., D614G, Alpha, Beta, Delta). In reality, a full schedule of vaccination likely confers lower level of protection against infection over long time scales due to waning, irrespective of the vaccine platforms *(15)*. Differences in long-term protection against onward transmission and hospitalization remain unclear. For simplicity and given the relatively short time scale being considered, we also assume that the first two antigen exposures dominate the acquisition of immunity and subsequent infections by Omicron beyond the first two may not change immune memory against Omicron *(26)*. An exhaustive list of all immunity protection scenarios considered in this study is shown in Table S3.

**Table S3:** List of the definitions of different types of protective immunity.

| Variable name | Definition | Value/Expression | Notes & Reference |
| --- | --- | --- | --- |
| $RR_{pom}^i(pOm)$ | Relative risk of infection by a pre-Omicron variant among individuals previously infected by a pre-Omicron variant, or in vaccinated individuals (breakthrough infection), compared to naïve individuals, and conditional on the same contacts. | 0.12 | Estimated from this study. |
| $RR_{pom}^{t\vert i}(pOm)$ | Relative risk of onward transmission of pre-Omicron variants among individuals infected with pre-Omicron variant for the second time, or in vaccinated individuals (breakthrough infection), compared to naïve individuals, and conditional on infection. | 0.4 | (17) |
| $\sigma_{om}^i$ | Degree of immune evasion with respect to susceptibility to infection by the Omicron variant. | NA | Free parameter to be explored in this study (Figure 4A). See also expression for $RR_{pom}^i(om)$. |
| $\sigma_{om}^{t\vert i}$ | Degree of immune evasion with respect to reduction in onward transmission by the Omicron variant, conditional on infection. | NA | Free parameter to be explored in this study (Figure 4A). |
| $\rho_{pom}^i$ | Degree of boosted immunity by pre-Omicron infection/vaccination with respect to reducing susceptibility to infection. | 0.5 | Assumed, see also expression for $RR_{pom-pom}^i(pOm)$ |
| $\rho_{pom}^{t\vert i}$ | Degree of boosted immunity by pre-Omicron infection/vaccination with respect to reduction in onward transmission. | 0.5 | Assumed. |
| $\rho_{pom}^{h\vert i}$ | Degree of boosted immunity by pre-Omicron infection/vaccination with respect to reduction in hospital admission. | 0.5 | Assumed. |
| $\rho_{om}^i$ | Degree of boosted immunity by Omicron infection/vaccination with respect to reduction in susceptibility to infection. | NA | Free parameter to be explored in this study (Figure 4D). |
| $\rho_{om}^{t\vert i}$ | Degree of boosted immunity by Omicron infection/vaccination with respect to reduction in onward transmission. | NA | Free parameter to be explored in this study (Figure 4D). |
| $RR_{pom}^i(om)$ | Relative risk of infection by Omicron strain between population with prior infection by pre-Omicron strain/full schedule of vaccination* and naïve population, conditional on the same exposure. | $\left(RR_{pom}^i(pOm)\right)^{1-\sigma_{om}^i}$ | |
| $RR_{pom}^{t\vert i}(om)$ | Relative risk of onward transmission by Omicron strain (given infection) between population with prior infection by pre-Omicron strain/full schedule of vaccination* and naïve population, conditional on infection. | $\left(RR_{pom}^{t\vert i}(pOm)\right)^{1-\sigma_{om}^{t\vert i}}$ | |
| $RR_{pOm-pOm}^i(pOm)$ | Relative risk of infection by a pre-Omicron strain in individuals with more than one prior infection/vaccination* (both primary and secondary infections are pre-Omicron), compared to naïve individuals. | $\left(RR_{pOm}^i(pOm)\right)^{1+\rho_{pOm}^i}$ | |
| $RR_{pOm-pOm}^{t\vert i}(pOm)$ | Relative risk of onward transmission (given infection by a pre-Omicron strain) in individuals with more than one prior infection/vaccination* (both primary and secondary infections are pre-Omicron), compared to naïve individuals. | $\left(RR_{pOm}^{t\vert i}(pOm)\right)^{1+\rho_{pOm}^{t\vert i}}$ | |
| $RR_{pOm-pOm}^i(Om)$ | Relative risk of infection by Omicron in individuals with more than one prior infection/vaccination* (both primary and secondary infections are pre-Omicron), compared to naïve individuals. | $\left(RR_{pOm}^i(pOm)\right)^{1+\rho_{pOm}^i-\sigma_{Om}^i}$ | |
| $RR_{pOm-pOm}^{t\vert i}(Om)$ | Relative risk of onward transmission (given infection by Omicron) in individuals with more than one prior infection/vaccination* (both primary and secondary infections are pre-Omicron), compared to naïve individuals. | $\left(RR_{pOm}^{t\vert i}(pOm)\right)^{1+\rho_{pOm}^{t\vert i}-\sigma_{Om}^{t\vert i}}$ | |
| $RR_{pOm-om}^i(Om)$ | Relative risk of infection by Omicron in individuals with more than one prior infection/vaccination* (with primary infection by a pre-Omicron strain/vaccination and secondary infection by Omicron), compared to naïve individuals. | $\left(RR_{pOm}^i(pOm)\right)^{1-\sigma_{Om}^i+\rho_{Om}^i}$ | |
| $RR_{pOm-om}^{t\vert i}(Om)$ | Relative risk of onward transmission (given infection by Omicron) in individuals with more than one prior infection/vaccination* (with primary infection by a pre-Omicron strain/vaccination and secondary infection by Omicron), compared to naïve individuals. | $\left(RR_{pOm}^{t\vert i}(pOm)\right)^{1-\sigma_{Om}^{t\vert i}+\rho_{Om}^{t\vert i}}$ | |
| $RR_{Om}^i(Om)$ | Relative risk of infection by Omicron strain in individuals with prior infection by Omicron, compared to naïve individuals, conditional on the same contacts. | $RR_{pOm}^i(pOm)$ | Assuming similar level of protection when compared to pre-Omicron strains |
| $RR_{Om}^{t\vert i}(Om)$ | Relative risk of onward transmission (given infection by Omicron) in individuals with prior infection with Omicron, compared to naïve individuals. | $RR_{pOm}^{t\vert i}(pOm)$ | Assuming similar level of protection when compared to pre-Omicron strains |
| $RR_{Om-om}^i(Om)$ | Relative risk of infection by Omicron in individuals with more than one prior infection (both primary and secondary infections are with Omicron), compared to naïve individuals. | $RR_{pOm-pOm}^i(pOm)$ | Assuming similar level of protection when compared to pre-Omicron strains |
| $RR_{Om-Om}^{ti}(Om)$ | Relative risk of onward transmission (given infection by Omicron) in individuals with more than one prior infection (both primary and secondary infections are with Omicron), compared to naïve individuals. | $RR_{pOm-pOm}^{ti}(pOm)$ | Assuming similar level of protection when compared to pre-Omicron strains |
| $RR_{Om-Om}^{hi}(Om)$ | Relative risk of hospital admission (given infection by Omicron) in individuals with more than one prior infection (both primary and secondary infections are with Omicron), compared to naïve individuals. | $RR_{pOm-pOm}^{hi}(pOm)$ | Assuming similar level of protection when compared to pre-Omicron strains |

\* For simplicity, given the low vaccination rate in South Africa, we assume that a full schedule of vaccination confers a similar level of protection as infection with pre-Omicron strains (i.e., D614G, Alpha, Beta, Delta). In reality, infection-induced immunity may confer superior protection against infection in the long run, relative to vaccination (20), while differences in magnitude and duration of protection against transmission and hospitalization remain unclear.

##### 5.4 Estimating the growth advantage of Omicron over Delta during its initial emergence

When Omicron was discovered in South Africa, the Delta epidemic had already declined and the Delta variant was circulating at low level in most locations *(20)*. In Figure S7, the dots show the logarithmic of SARS-CoV-2 weekly incidence in the District of Kenneth Kaunda between weeks 35 and 48 in 2021. The epidemic curve can be viewed as the supposition of an exponential decay and an exponential growth, with the transition occurring around week 45 of 2021 (coinciding with the emergence of Omicron in South Africa). Here we assume that the exponential decay (prior to week 45, 2021) was driven by the Delta variant and the exponential growth (post week 45, 2021) was driven by the Omicron variant. We can thus model the epidemic curve as supposition of exponential decay and exponential growth, i.e.

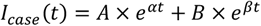

Here α is the growth rate of Delta and α is the growth rate of Omicron and *A* and*B* are the initial Incidence rate for Delta and Omicron when *t* = 0. We fit this function to the observed epidemic curve between week 35 and week 48 using maximum likelihood method. We find a growth rate of -0.063 per day for the Delta variant (exponential decay) and 0.275 per day for the Omicron variant (exponential growth), indicating a growth advantage of 0.338 per day of Omicron over Delta in Dr Kenneth Kaunda District. Figure S7 shows the results of the fitting.

##### 5.5 State-space transmission model for Delta variant and projection of Delta spread from weeks 35 to 45, 2021

Here we consider a “Susceptible-Infectious-Recovered-Susceptible” (SIRS) model for Delta that that tracks infection history up to 3 repeat infections/immunizations. The equations governing the model are as follows:

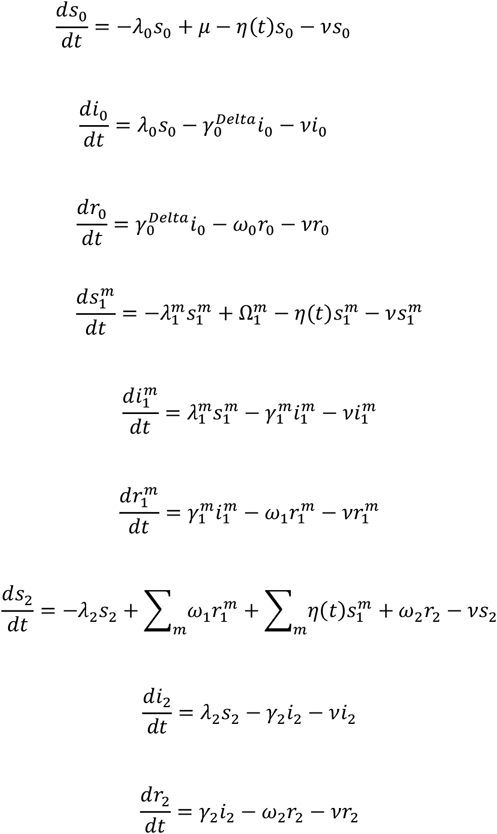

With the following expression for the force-of-infection:

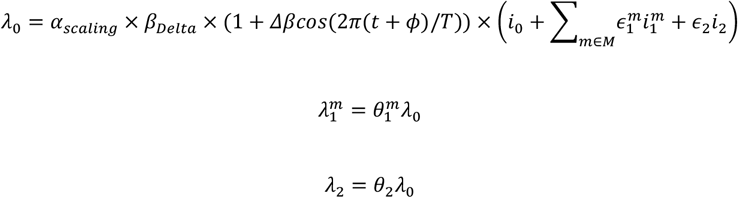

The infectious periods for 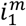 and *i*_2_ can be expressed as follows with respect to *i*_0_’s:

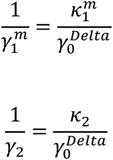

The definitions of the state variables are presented in Table S4, and the definitions of the model parameters are in Table S5. We initialized the model based on the reconstructed exposure history (Figure S6) by week 34. The only free parameter of the model was the rescaling factor on transmissibility *α*_*scaling*_ (Table S5). We optimized *α*_*scaling*_ so that the projected curve of new incidence would decay at a rate of -0.063 day^-1^, matching the observation (Figure S7). The best fit for *α*_*scaling*_ was 0.44. We further projected the SARS-CoV-2 antigen exposure history from week 35 to week 45 (prior to the emergence of the Omicron variant (Figure S8).

**Table S4:** State variables of the Delta variant compartmental transmission model.

| Variable name | Variable type | Definition |
| --- | --- | --- |
| $m \in M$ | Categorical | $m$ Indicates the type of primary SARS-CoV-2 antigen exposure from the set $M = \{D614G, Beta, Delta, Delta, Others, Vacc\}$ denotes immune history including only one antigen exposure by pre-Omicron infection (including D614G, Beta, Delta, and other variants other than Omicron (“Others”) in South Africa) or a primary schedule of vaccination (“Vacc”). |
| $s_0$ | State variable | Fraction of susceptible in the population who are fully naïve against any SARS-CoV-2 infection and are unvaccinated. |
| $s_1^m$ | State variable | Fraction of susceptible in the population who have experienced one infection or a full schedule of vaccination, with $m$ denoting the type of primary antigen exposure (i.e., variant type if infection or if primed by vaccination). |
| $s_2$ | State variable | Fraction of susceptible in the population who have experienced two or more infections and immunization combined. We assume that the first two immunizations will have the strongest impact on the level of long-term protective immunity. |
| $i_0$ | State variable | Fraction of population who got infected by Delta from population in $s_0$ |
| $i_1^m$ | State variable | Fraction of population who got infected by Delta from population in $s_1^m$ |
| $i_2$ | State variable | Fraction of population who got infected by Delta from population in $s_2$ |
| $r_0$ | State variable | Fraction of population who recovers from Delta in $i_0$ compartment and enjoys a temporary period of full immunity (100% protection) against any reinfection. |
| $r_1^m$ | State variable | Fraction of population who recovers from reinfection/breakthrough of Delta infection in $i_1^m$ compartment and enjoys a temporary period of full immunity (100% protection) against any reinfection. |
| $r_2$ | State variable | Fraction of population who recovers from further reinfection/breakthrough of Delta infection in the $i_2$ compartment and enjoys a temporary period of full immunity (100% protection) against any reinfection. |

**Table S5:**
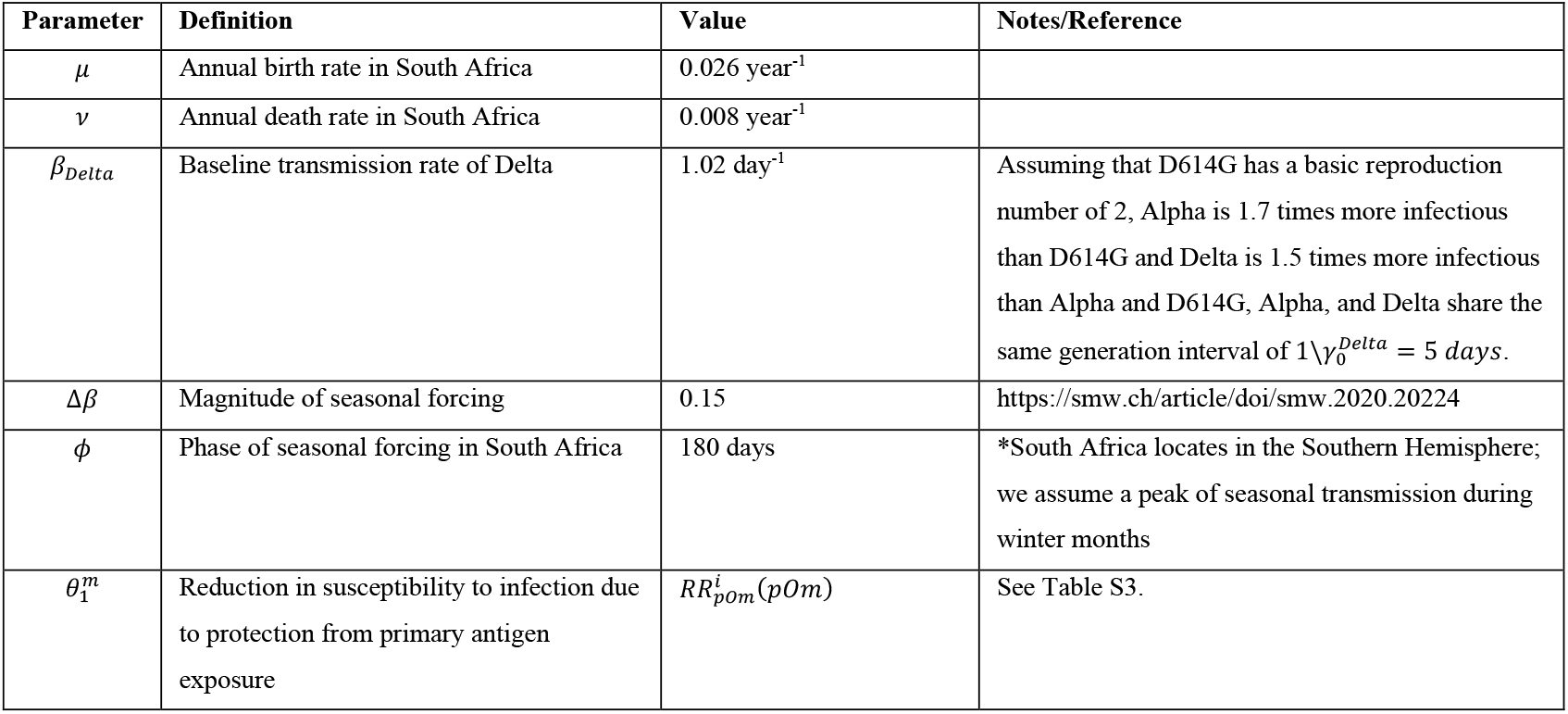
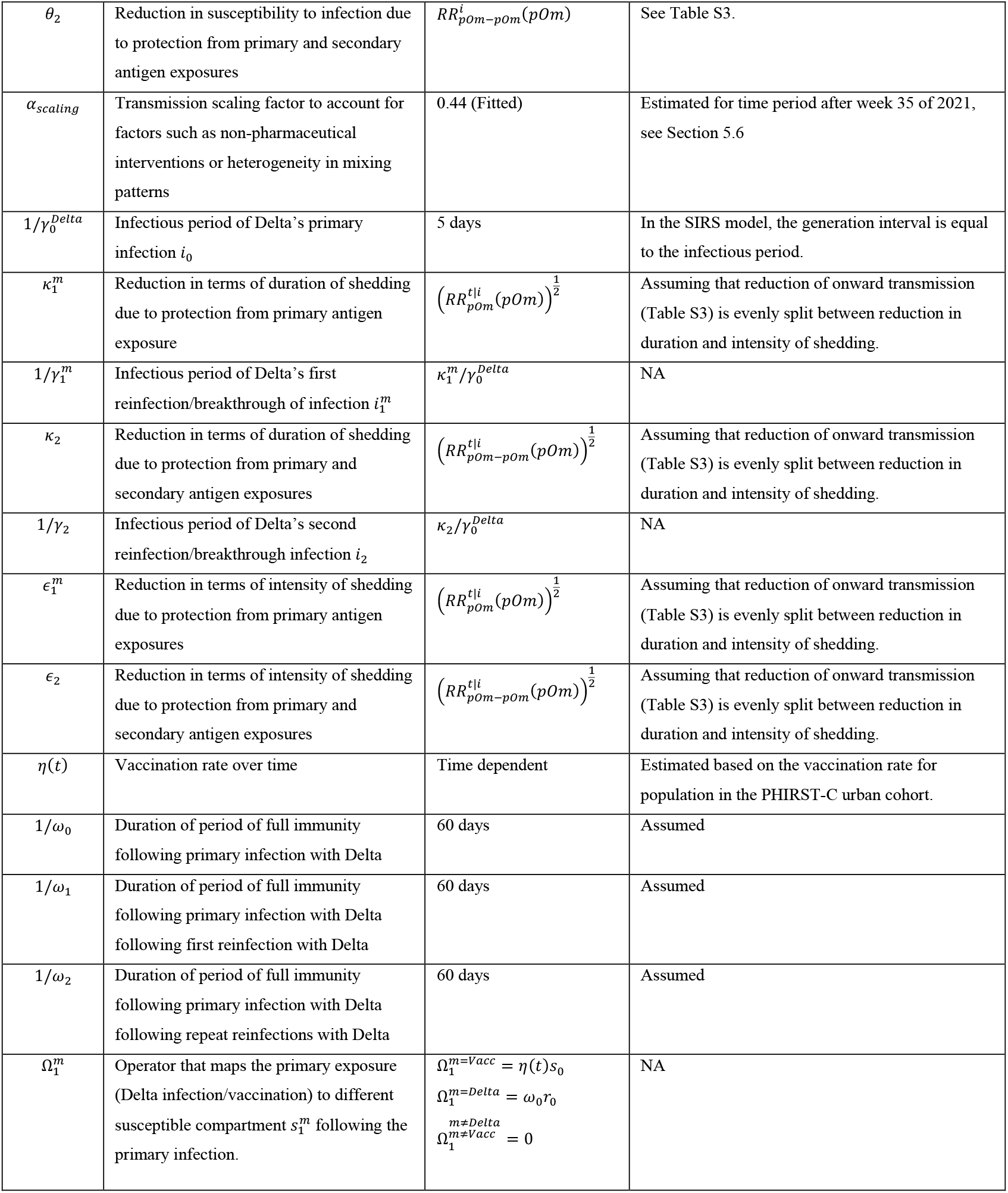
Model parameters for the Delta variant transmission model.

##### 5.6 State-space transmission model for the Omicron variant and projection of Omicron spread from week 45, 2021 to the end of the Omicron wave

For simplicity, we consider a hypothetical scenario where Omicron has successfully displaced all other circulating variants in South Africa and explore how the transmission dynamics of Omicron is shaped by the immune history of previously circulating strains and vaccination. Accordingly, at the time of writing, Omicron had replaced Delta in many countries that report variant-specific prevalence estimates, including South Africa. We do not consider variant co-circulation and the emergence of new variant during the Omicron wave, although such scenarios are certainly possible. Similar to the Delta variant, we consider a “Susceptible-Infectious-Recovered-Susceptible” model for Omicron that tracks infection history up to 3 repeat infections/immunizations, with additional Omicron-specific properties of immune evasion and enhanced transmissibility. The equations governing the model are as follows:

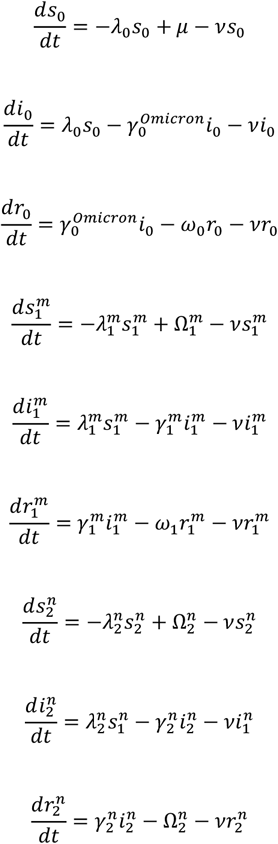

With the following expression for the force-of-infection:

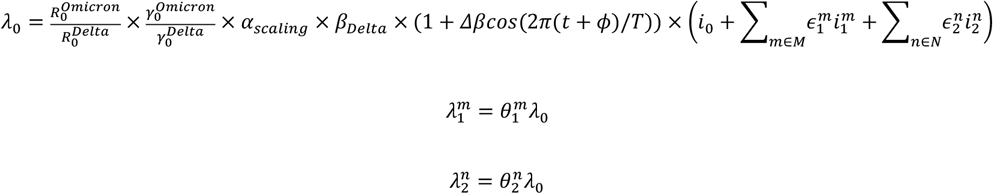

The infectious periods for 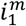 and 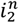 can be expressed as follows with respect to *i*_0_ ‘s:

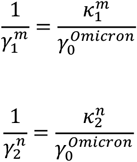

The definitions of the state variables are presented in Table S6, and the definitions of the model parameters are in Table S7.

Based on the transmission model, we explored how the degree of Omicron’s immune evasion against infection 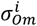 and onward transmission 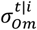 would shape the trajectory of the epidemic (See Section 5.2 and Table S3 for the definition of 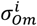 and 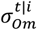 We scanned through values of 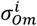 and 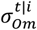 ranging from 0 to 1, with a step size of 1/30. We also considered a potential change in the generation interval (GI) of Omicron when compared to the Delta variant. We explored possible values for Omicron’s generation time, including 3, 4, 5, and 6 days. For each pair value pair of 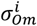 and 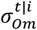 and GI, we fit the ratio of basic reproduction number between Omicron and Delta 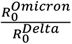 so that the growth rate of Omicron matched the observed initial growth of the Omicron wave (Figure S7). We fit the fraction of individual in the *i*_0_ compartment at week 45, 2021 so that the peak of the projected incidence of Omicron infections matched the observed Omicron case incidence.

We then calculated the characteristics of the projected Omicron wave, including the estimated 1) infection attack rate, 2) epidemic duration, 3) fraction of reinfections/breakthrough infections among all infections, 4) the relative reduction of realized GI (average GI over both primary infections and reinfections/breakthrough of infections) with respect to intrinsic GI, and the 5) infection case ratio (number of cases reported to the Dr Kenneth Kaunda District during the Omicron wave divided by the total number of projected Omicron infections), for a given 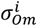 and 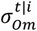 and GI.

Figure S9 visualizes projected characteristics of the Omicron wave as a function of 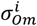 and 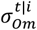 and GI. Figure 4A-E represents a scenario where the GI of Omicron is 4 days, shorter than Delta *(11, 17, 18, 26)*, where white dots represent our best knowledge of the degree of Omicron’s evasion of prior immunity against infection and onward transmission *(64)*.

**Table S6:**
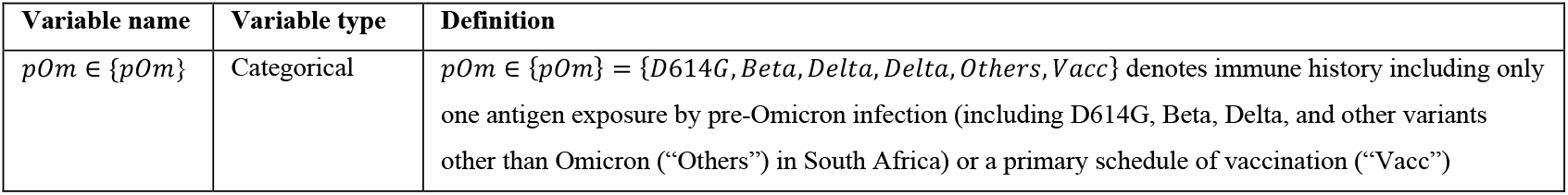
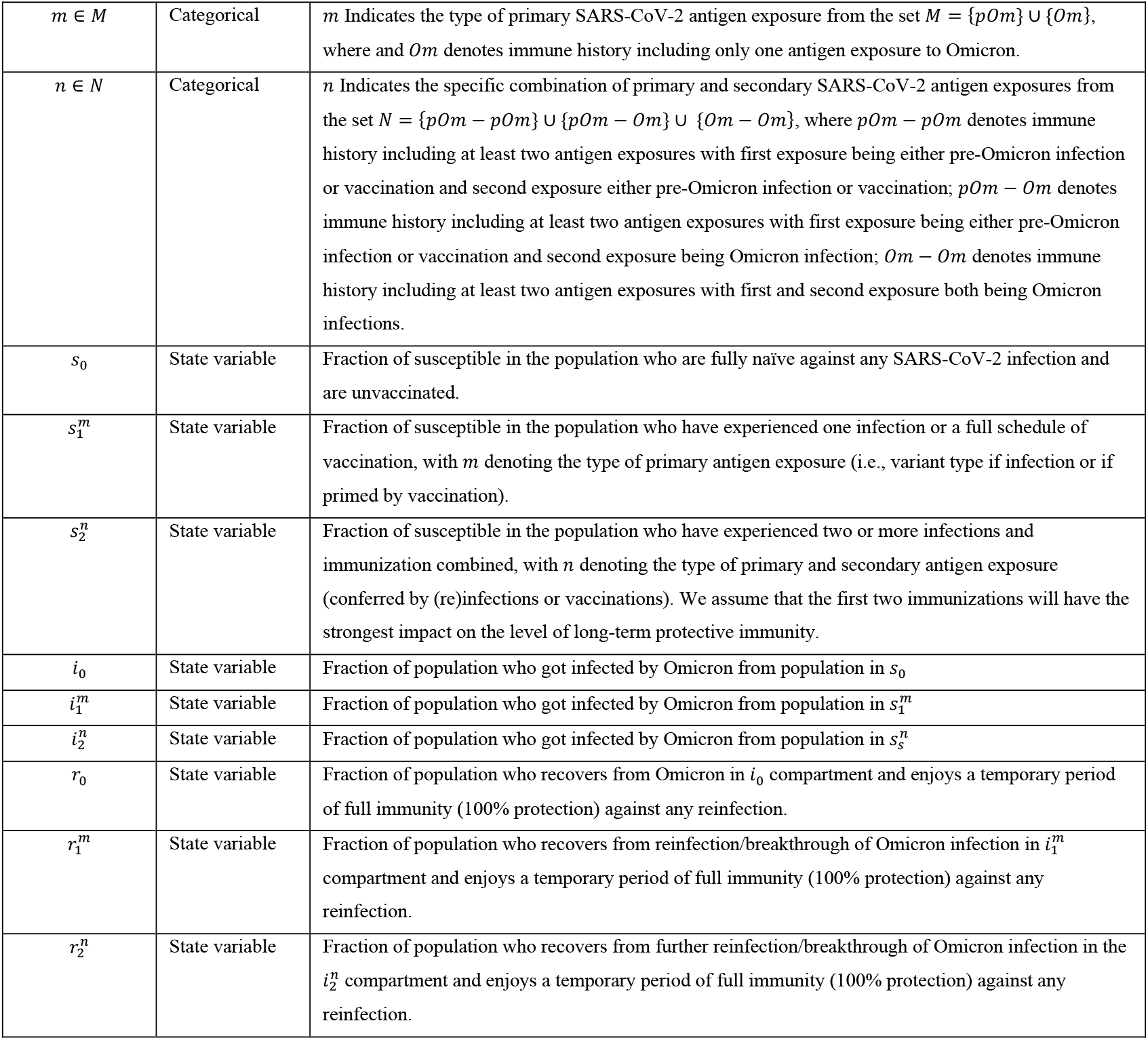
State variables of the Omicron variant compartmental transmission model.

**Table S7:**
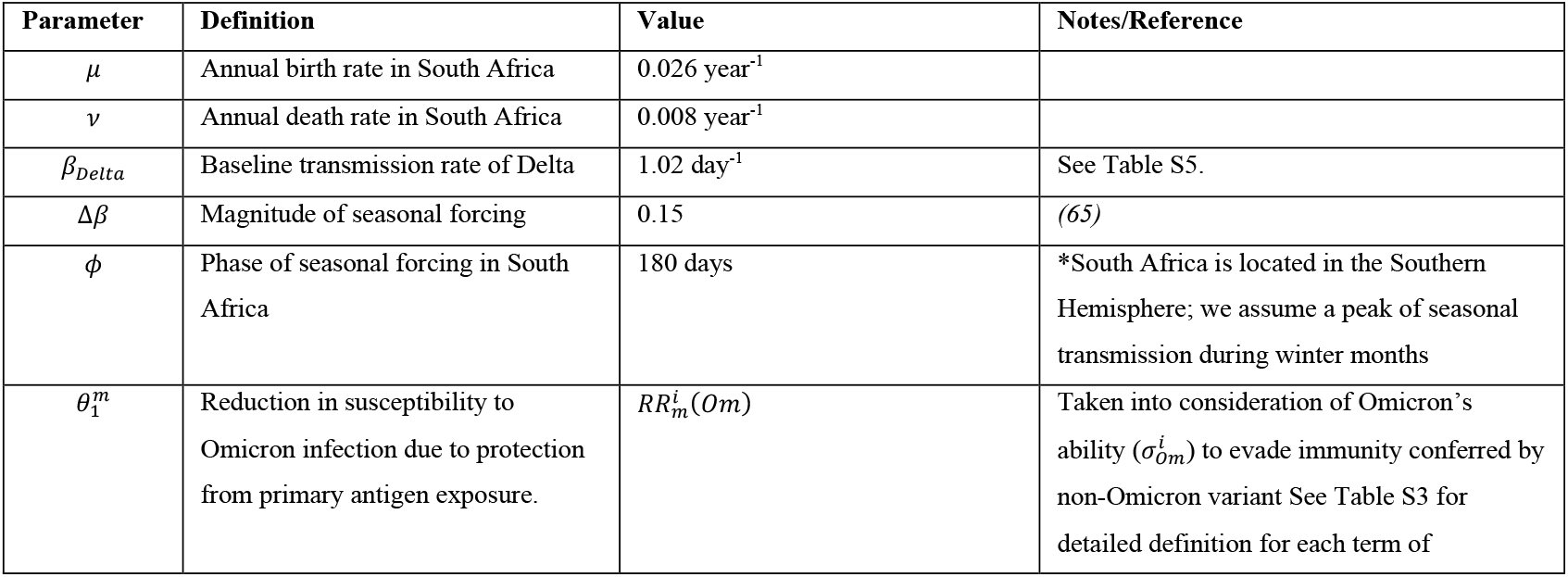
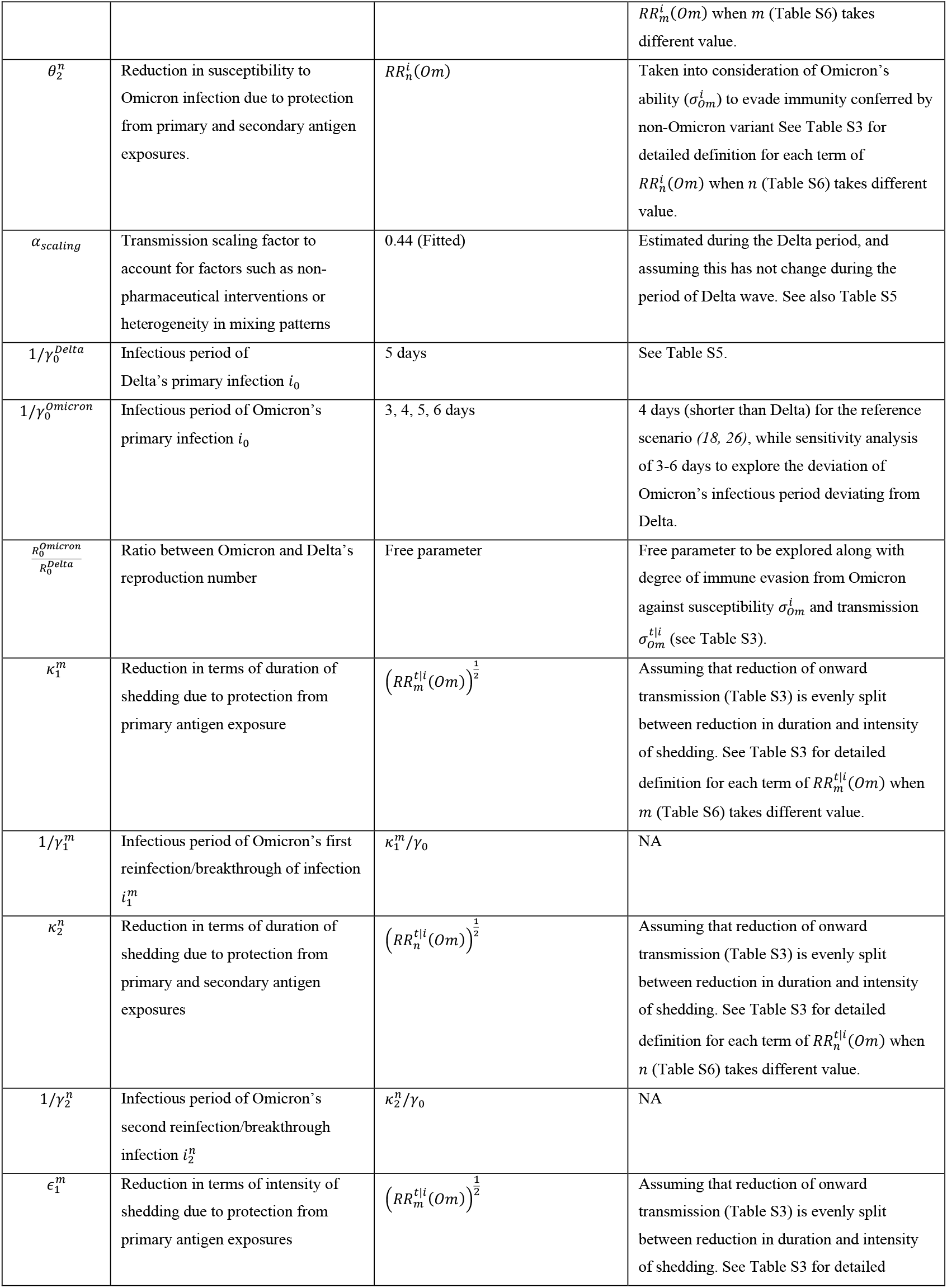
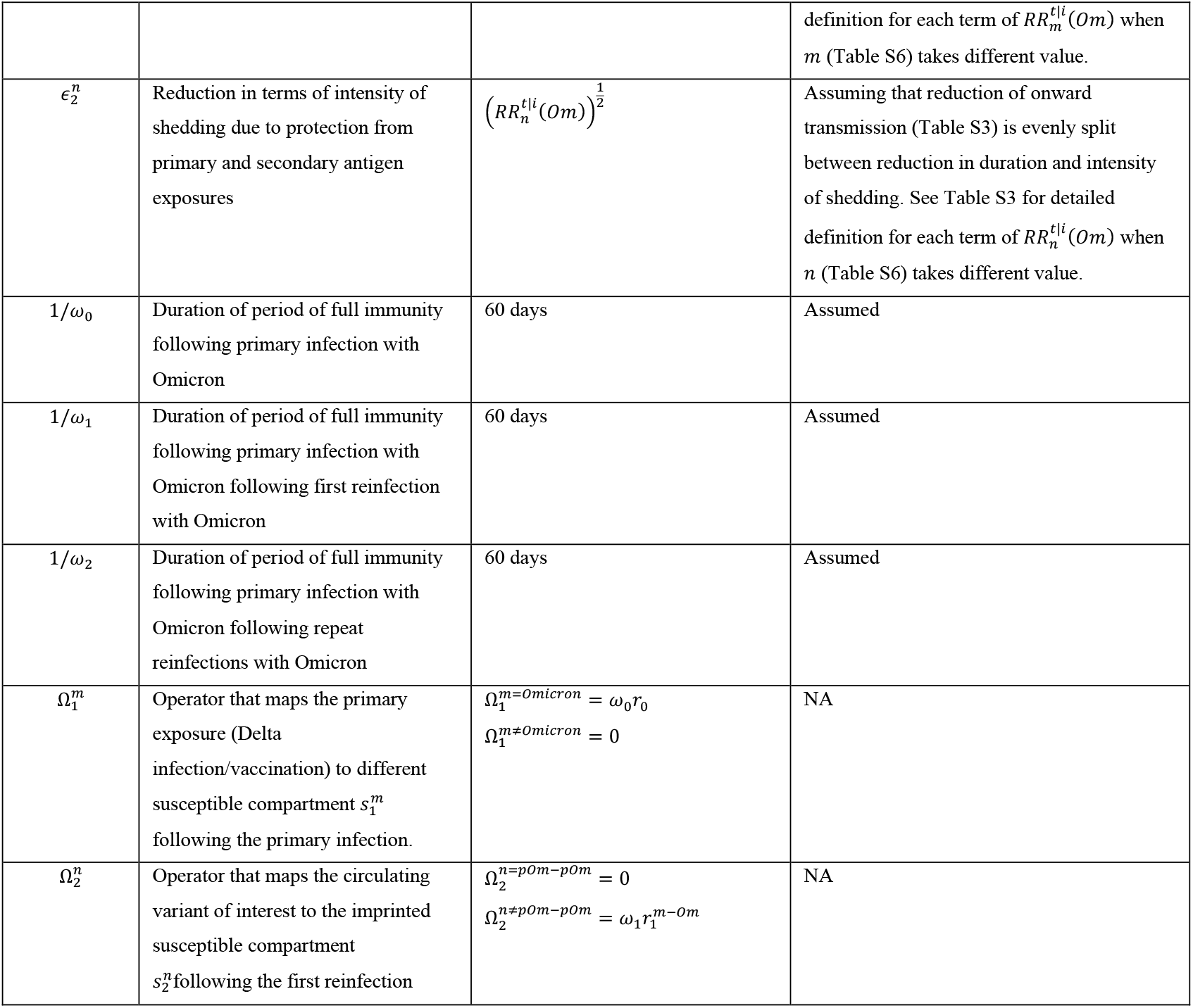
Model parameters for the Omicron variant transmission model.

#### 6. Modelling the transmission dynamics of Omicron, Delta and a hypothetical variant X after the Omicron wave

Here we modify the transmission model described in Section 5 to evaluate the possibility of a fifth epidemic wave after the Omicron wave. Specifically, we evaluate the potential recurrence of three variants independently: Omicron, Delta, and a hypothetical variant X, where X is at equal antigenic distance from Omicron and Delta. We consider the projected Omicron wave for the reference scenario (RS) shown in Figure 4F. After the Omicron wave, two new population groups need to be taken into consideration based on their antigen exposure(s): 1) individuals who were primed by the Omicron variant, which accounts for 23% of the population; and 2) individuals who were primed by a non-Omicron variant (through infection or vaccination) then reinfected by Omicron, which accounts for 46% of the population. For any given variant of interest, we denote the level of protection (against infection) conferred after primary Omicron infection as 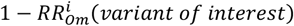 and after Omicron reinfection/vaccine breakthrough as 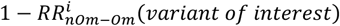, where as *RR*^*i*^ stands for the relative risk of acquiring infection when compared to an immunologically naïve individual (1 − *RR*^*i*^ = 0% indicates no protection while 1 − *RR*^*i*^ = 100% indicates perfect protection). For simplicity, we assume protection against transmission 1 − *RR*^*t* |*i*^ remains constant at 60% *(18, 26)*.

##### 6.1 Omicron transmission model

For the Omicron transmission model, we first consider the same parameters as in the reference scenario as shown in Figure 4F, with generation time of 4 days, 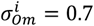, 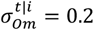, and the estimated 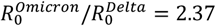. However, for the two population groups of interest 1) those who were primed by Omicron 2) those who have experienced a Omicron reinfection/breakthrough we consider their relative risk (with respect to naïve population) of acquiring Omicron infection as 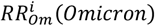 and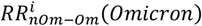, respectively. In Figure S10A, we use the transmission model to evaluate the growth rate of Omicron when Omicron is reintroduced into the population for all combinations of 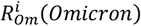 and 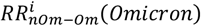 ranging from 0 to 1. A growth rate larger than 0 indicates that the Omicron variant is above the epidemic threshold, leading to a recurring fifth epidemic wave, while a growth rate lower than 0 indicates that the Omicron variant will not trigger another outbreak after the fourth wave. We further consider a scenario (Figure S10B) where the contact rate is twice that of the one during the fourth wave, i.e., *α*_*scaling*_= 0.44 × 2 = 0.88.

##### 6.2 Delta transmission model

To evaluate the risk of Delta recurrence, we consider the same Delta transmission model described in Materials and Methods Section 5.5. However, for the population groups 1) who were primed by Omicron 2) who have experienced a Omicron reinfection/vaccine breakthrough, we consider their relative risk (with respect to naïve individuals) of acquiring Delta infection as 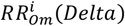 and 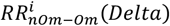, respectively. In Figure S10C, we use the transmission model to evaluate the growth rate of the Delta variant when it is reintroduced into the population for any combination of 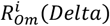 and 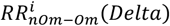 ranging from 0 to 1. We further consider a scenario (Figure S10D) where the contact rate is twice that of the one during the fourth wave, i.e., *α*_*scaling*_ = 0.44 × 2 = 0.88 (Table S5).

##### 6.3 Transmission model of the hypothetical variant X

For the transmission model of a hypothetical new variant X (Figure S10E), we consider that variant X has the same basic reproduction number and generation time as the Delta variant, and the contact rate is twice that of the fourth wave, i.e., *α*_*scaling*_ = 0.44 × 2 = 0.88. We additionally assume that variant X is antigenically equally distinct from both Omicron and pre-Omicron variant so that the relative risks of reinfection are equal irrespective of the primed strain i.e.,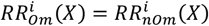. The rest of the model is the same as for Delta.

**Fig. S1:**
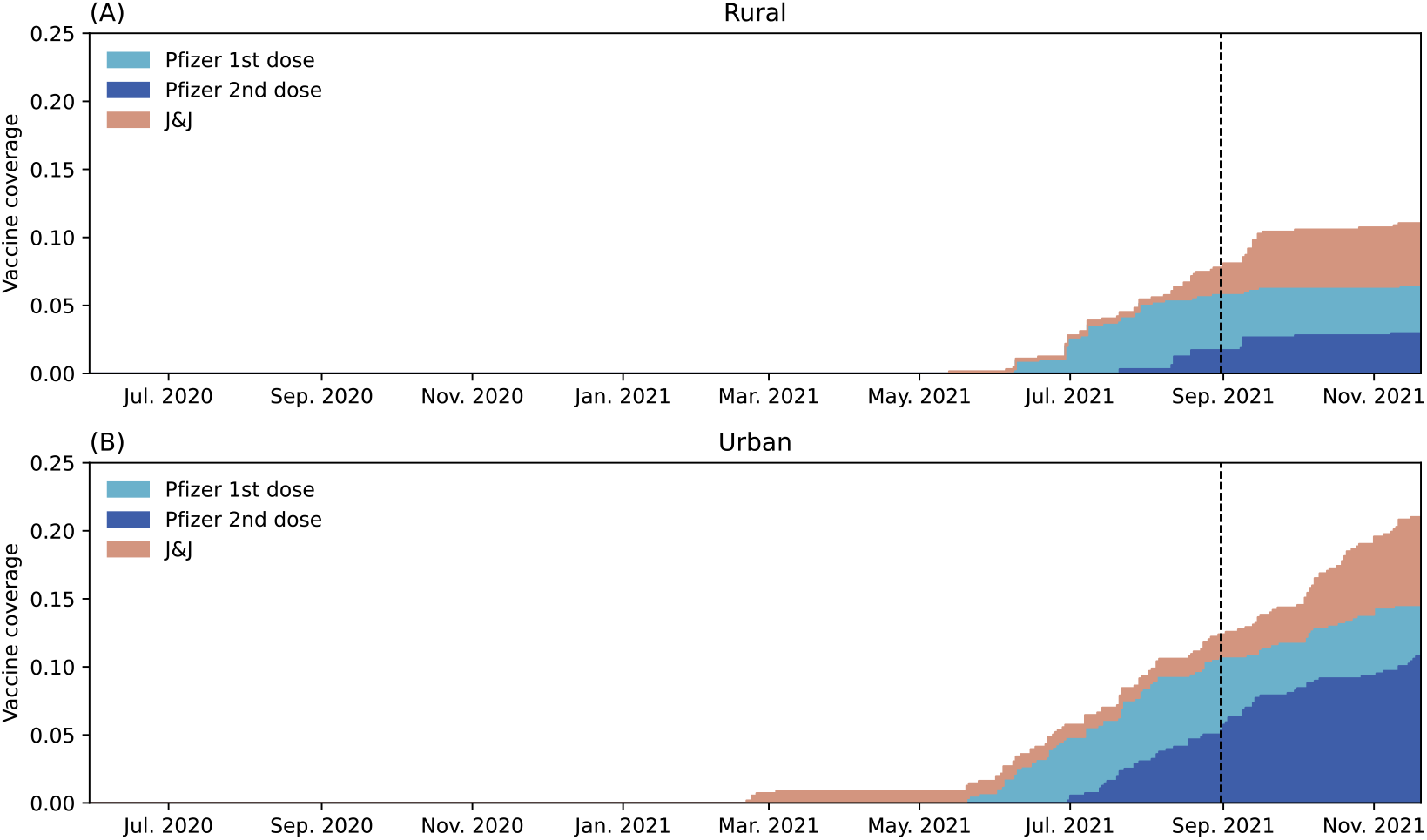
Vaccination rate by vaccine types, including J&J/Janssen Ad26.COV2.S (J&J) and the Pfizer/BioNTech BNT162b2 (Pfizer). The dashed lines indicate the end of PHIRT-C (August 28, 2021).

**Fig. S2:**
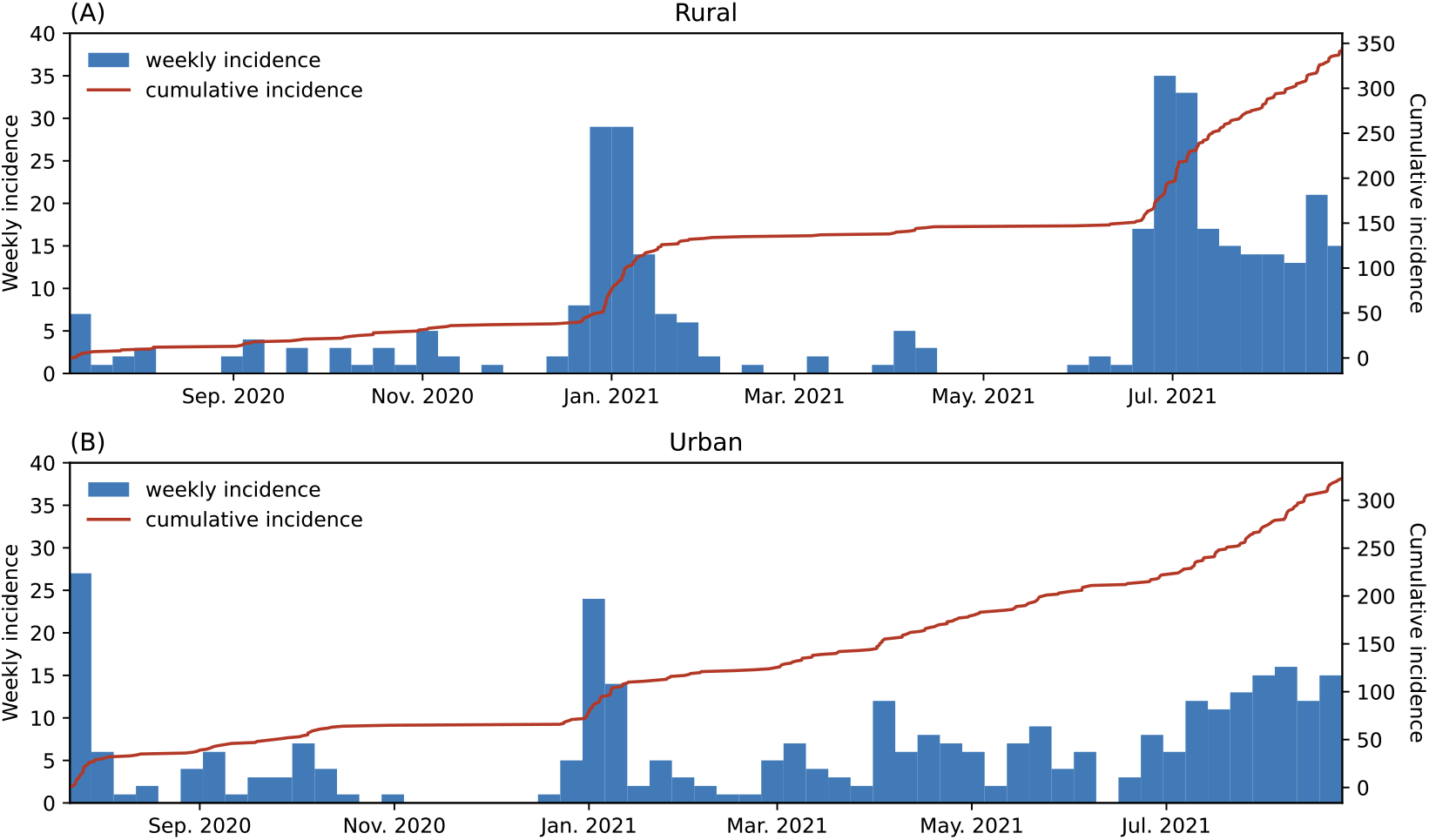
(A) Weekly incidence (blue bar) and cumulative incidence (red line) of SARS-CoV-2 infection among participants in the rural cohort. (B) Same as (A) but in the urban cohort.

**Fig. S3:**
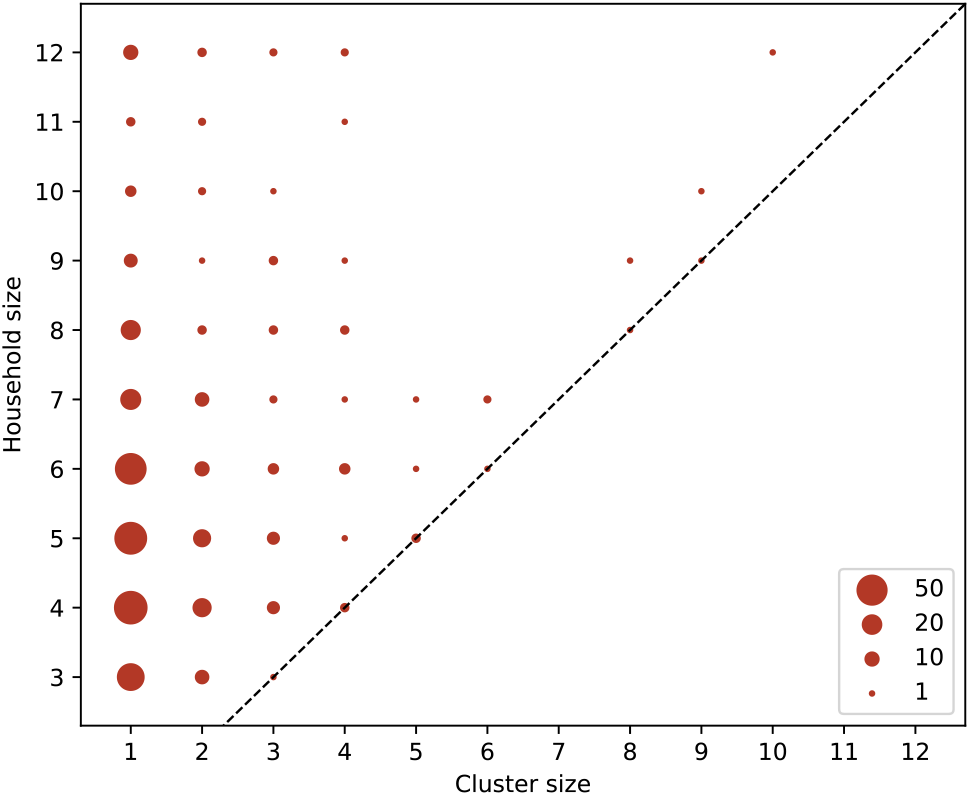
Distribution of the size of the household infection cluster at different household sizes across 222 households in both the rural and urban cohort. A household infection cluster with size larger than one is defined as a group of infections within the same household with at least two infection episodes within the same cluster with infection time separated no more than 14 days. An isolated infection episode is considered an infection cluster with cluster size one. The sizes of the dots are proportional to the frequency of occurrence for each “household size – cluster size” pair among 192 infection clusters.

**Fig. S4:**
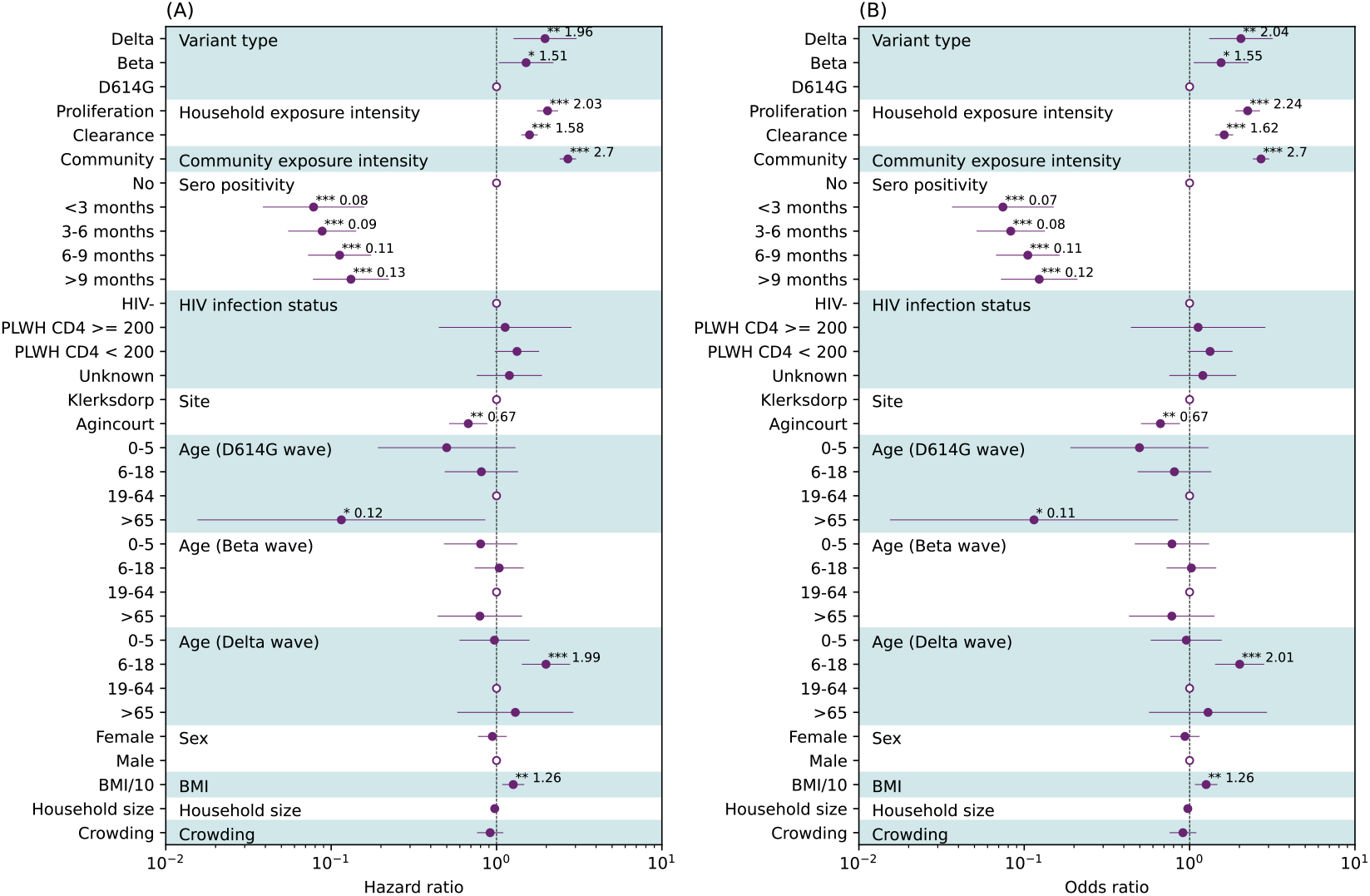
(A) Discrete time survival analysis using piecewise exponential model (Poisson regression). Hazard ratios (HR) along with 95%CIs are reported as solid dots and horizontal lines. The hollow dots are reference class for each of the categorical variable. Comparing to results presented in Figure 3C, we consider a censoring time window from the time of infection to 15 days after viral RNA clearance for each infection episode. (B) Discrete time survival analysis using piecewise logistic model (logistic regression). Comparing to results presented in Figure 3C, we consider the piecewise logistic model rather than Poisson model, with the same censoring time window. Odds ratios (OR) along with 95%CIs are reported as solid dots and horizontal lines. The hollow dots are reference class for each of the categorical variable. *Indicates p<0.05; ** indicates p<0.01; *** indicates p < 0.001. Abbreviations: HIV- (HIV-uninfected individuals), PLWH+ CD4 <200 (Persons living with HIV, CD4+ T cell count under 200 cells/ml), PLWH+ CD4 >=200 (Persons living with HIV, CD4+ T cell count equal or above 200 cells/ml).

**Fig. S5:**
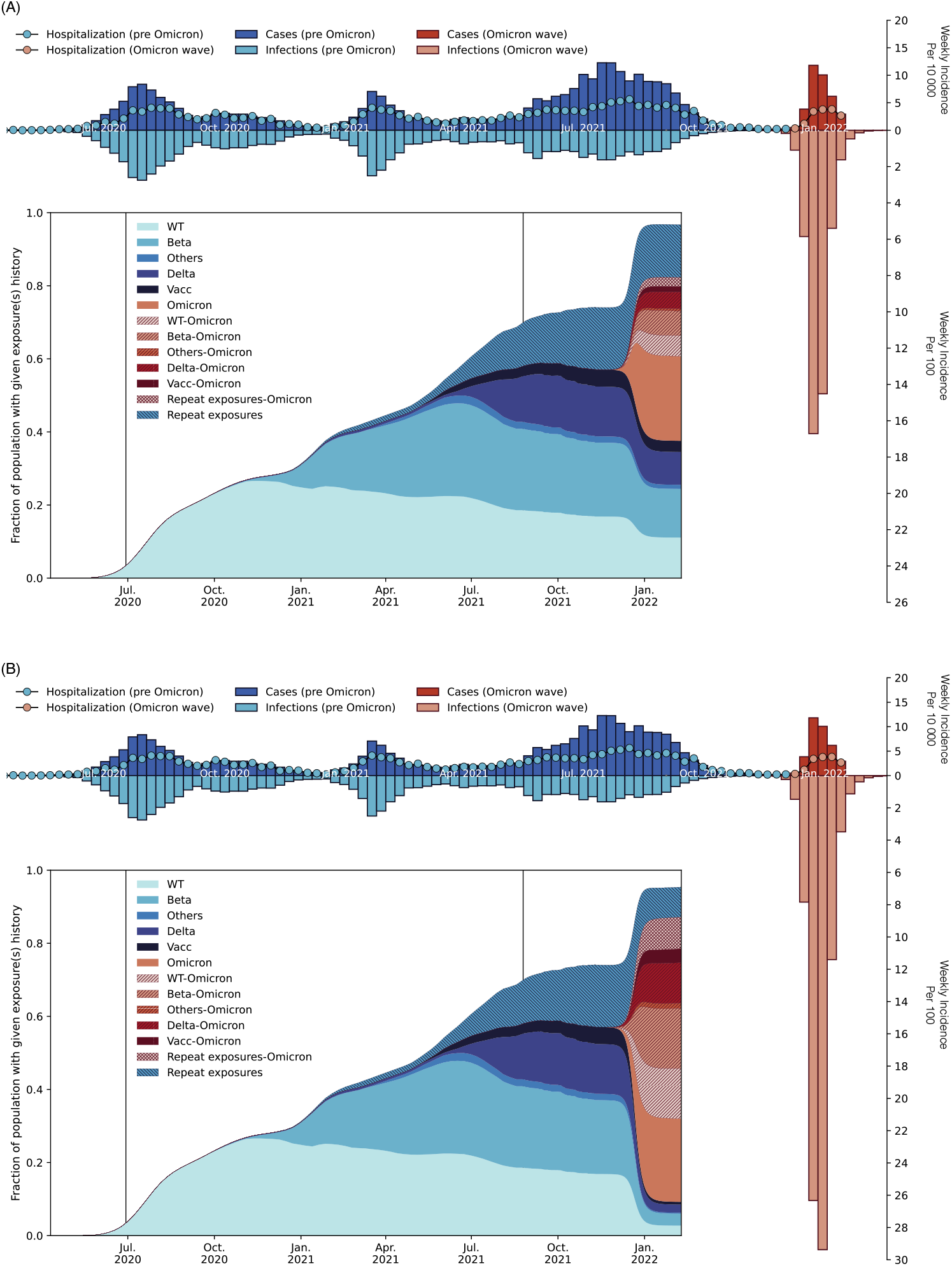
(A) Same as Figure 4F but for a low immune escape (LE) scenario with 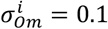 and 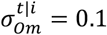. (B) Same as Figure 4F but for a high immune escape (HE) scenario with 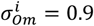 and 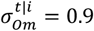.

**Fig. S6:**
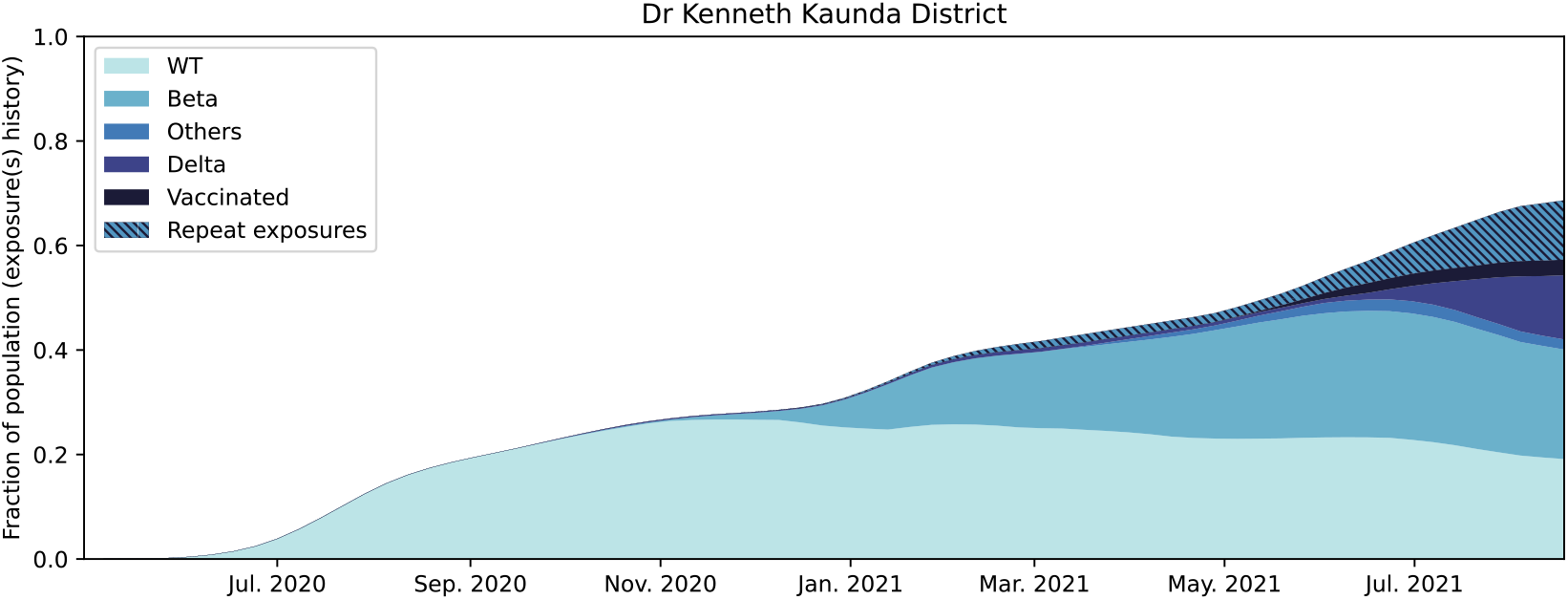
SARS-CoV-2 antigen exposure history by the end of PHIRST-C (September 2021) at the District of Dr Kenneth Kaunda.

**Fig. S7:**
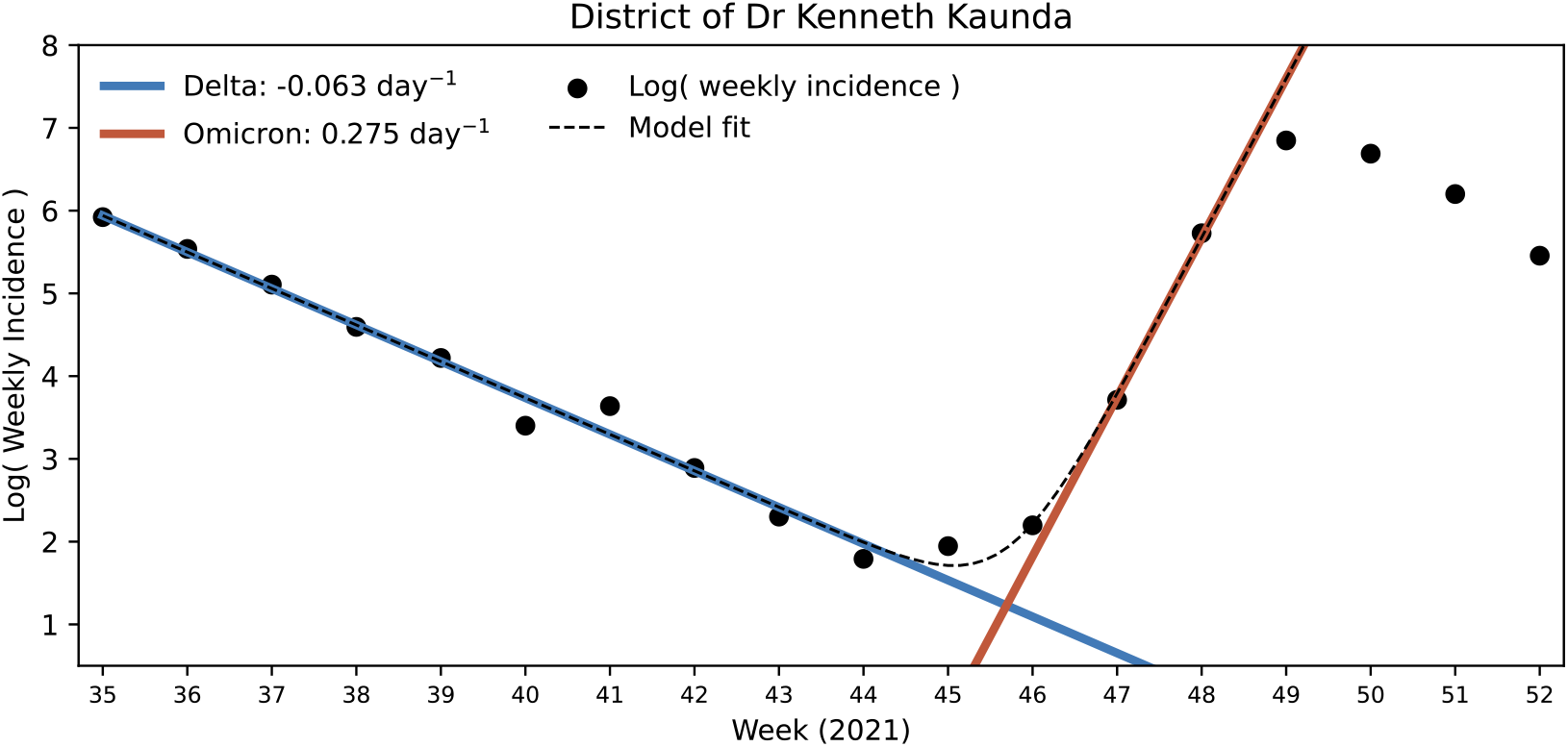
Estimated case growth rate of Delta and Omicron between weeks 35 and 52 of 2021. Dots are the logarithmic of the weekly incidence of SARS-CoV-2 cases reported to the District of Dr Kenneth Kaunda between weeks 35 and 52 of 2021. The blue line is the fitted exponential decaying of Delta Incidence while the red line is the fitted exponential growth of Omicron wave and the dashed line is the fitted convolution of the two variants.

**Fig. S8:**
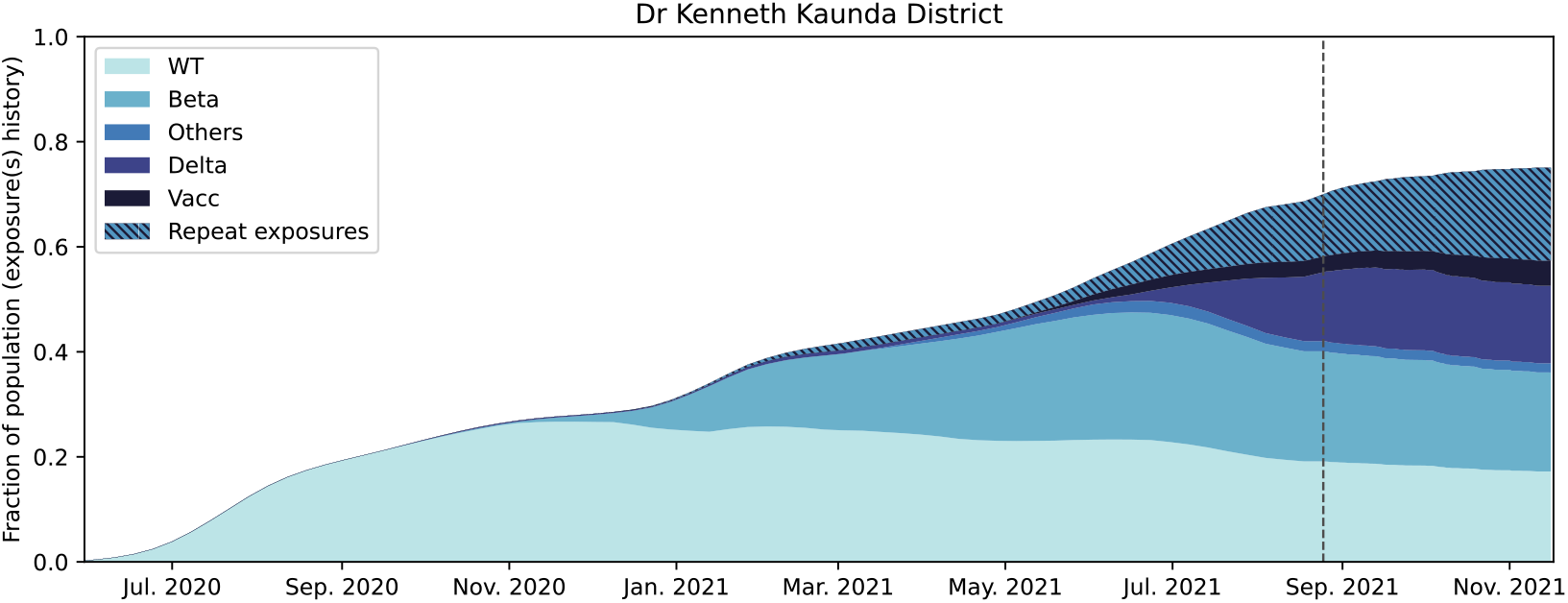
Projection of SARS-CoV-2 antigen exposure history from the end of PHIRST-C (week 35 of 2021, dashed line) and until the emergence of Omicron (week 45 of 2021, Figure S7) at the District of Dr Kenneth Kaunda.

**Fig. S9:**
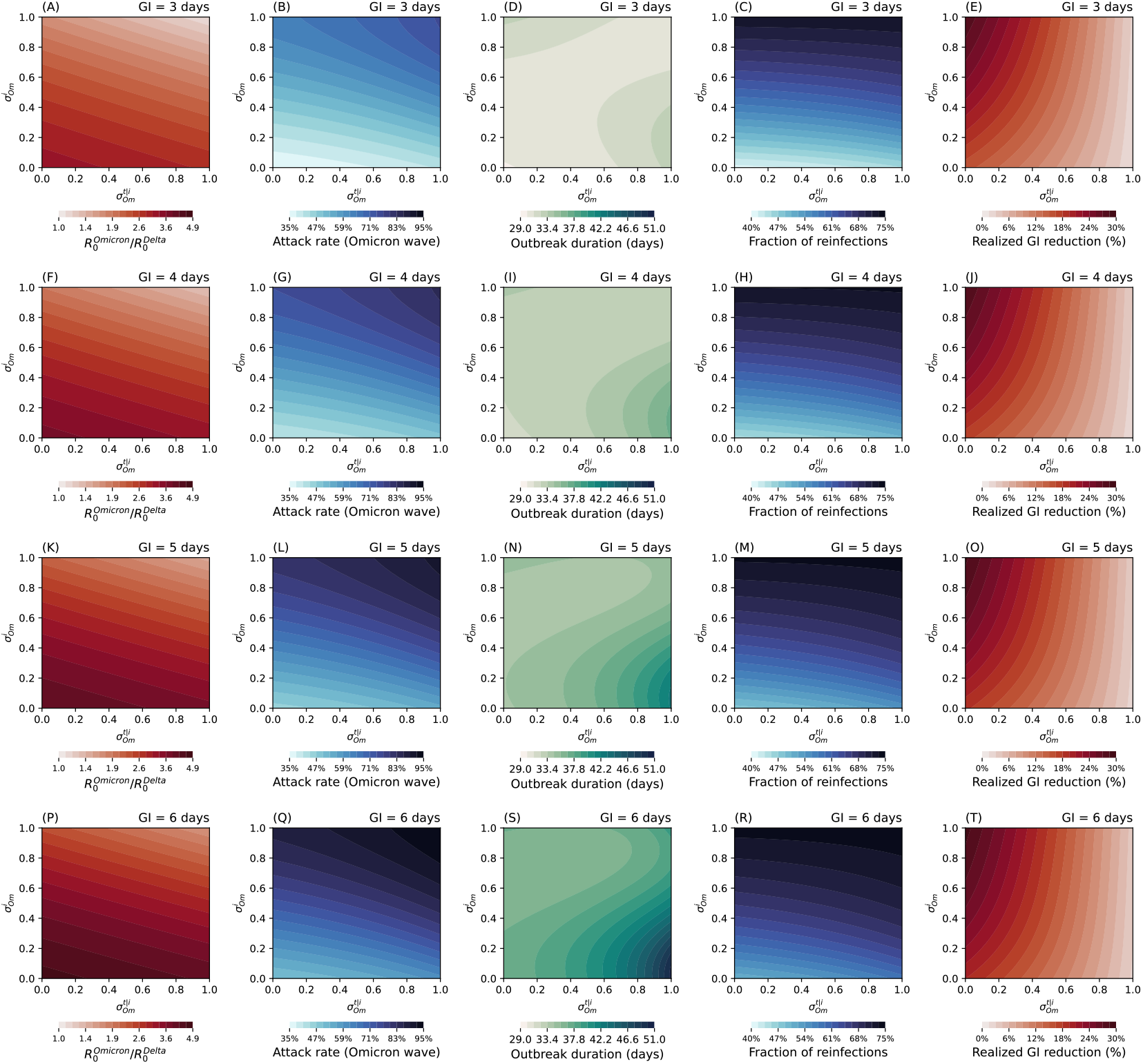
The characteristics of the Omicron epidemic wave as a function of 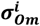 and 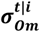and generation time. Each row corresponds to different values of generation time (GI) of primary infection. Column one corresponds to ratio of basic reproduction number between Omicron and Delta. Column two corresponds to infection attack rate. Column three corresponds to duration of the epidemic. Column four corresponds to fraction of reinfections/breakthrough of infections among all infections. Column five corresponds to the relative reduction of realized GI (average GI over both primary infections and reinfections/breakthrough infections) with respect to the intrinsic GI.

**Fig. S10:**
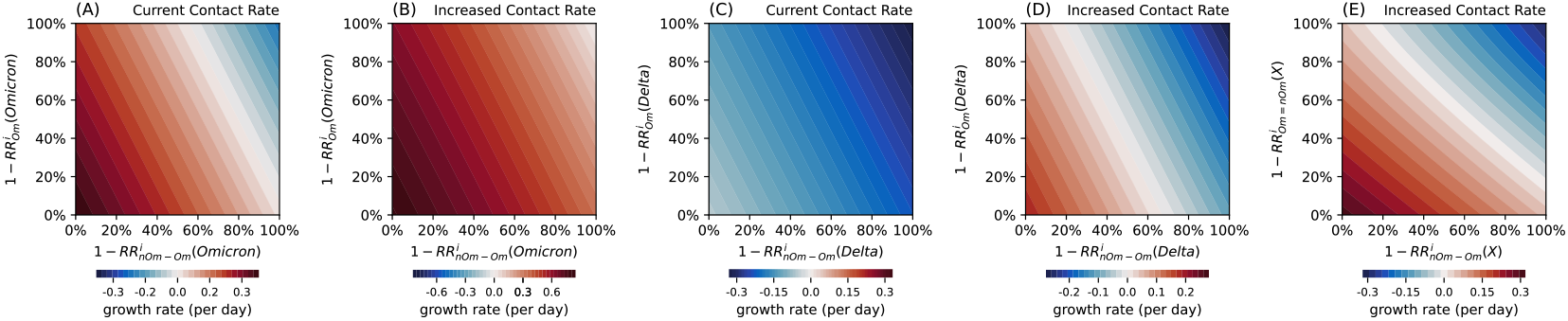
Possible post-Omicron futures, exploring potential resurgences of Omicron, Delta, or a new variant X, under different contact scenarios. Projections are based on the reconstructed immune histories at the end of January 2022 shown in Fig 4F (i.e., given a reference scenario for Omicron’s immune escape 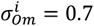 and 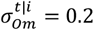). (A) Risk of recurrence of Omicron: phase diagram of the growth rate of Omicron in a recurrent wave, as a function of the level of protection conferred by Omicron primary infections against Omicron 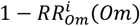, and the level of protection conferred by Omicron reinfections/breakthroughs against Omicron 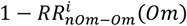. Contact rates are assumed to remain the same as during the Omicron wave. (B) same as (A) but assuming the contact rate is twice of that during the Omicron wave. (C) Risk of recurrence of Delta: Phase diagram of the growth rate of Delta, after the initial Omicron wave has subsided, as a function of the level of protection conferred by Omicron primary infection against Delta 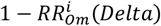, and the level of protection conferred by Omicron reinfections/breakthroughs against Delta 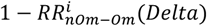. Contact rates are assumed to remain the same as during the Omicron wave. (D) same as (C) but assuming the contact rate is twice of that during the Omicron wave. (K) Risk of occurrence of hypothetical new variant X, where X is at equal antigenic distance of Delta and Omicron: Phase diagram of the growth rate of variant X, after the initial Omicron wave has subsided, as a function of the level of protection conferred by any variant primary infection on infection with 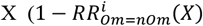, assuming Omicron and pre-Omicron infections confer the same level of protection against X), and the level of protection conferred by Omicron reinfections/breakthroughs on X infection,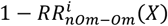. Contact rate is assumed to be twice of that of the Omicron wave.

